# Machine Learning Models Using Electronic Health Record Data to Predict Obstructive Sleep Apnea

**DOI:** 10.64898/2026.10.05.26364806

**Authors:** Nathanael H. Hwang, Diego R. Mazzotti, Soniya Mishra, M. Brandon Westover, Dennis Hwang, Robert J. Thomas, Emmanuel Mignot, Umakanth Katwa, Jiaxiao Shi, Rui Yan, Kendra A. Becker, Matthew T. Klimper, Jessica Jara, Damien R. Stevens, Matthew K.P. Gratton, Haoqi Sun

## Abstract

**Rationale:** Obstructive sleep apnea (OSA) is highly prevalent yet largely under-diagnosed. Current screening strategies rely on effortful questionnaires with modest accuracy, and prior machine learning models often require resource-intensive inputs and lack external validation.

**Objectives:** To develop and validate machine learning models to predict OSA (apnea-hypopnea index [AHI4%]≥5) and moderate-severe OSA (AHI4%≥15) using only routinely collected electronic health record (EHR) data, test cross-system transferability and compare performance with clinical screeners and a Bayesian risk-updating framework.

**Methods:** Three tree-ensemble families (XGBoost, random forests, and histogram-based gradient boosting) were trained on an adult cohort with sleep-study results at Kaiser Permanente (KPSC), using full (106-feature) and minimal (age, sex, body-mass index, race/ethnicity) predictor sets and tuned by nested cross-validation. XGBoost models were selected, internally and temporally validated, and externally validated at University of Kansas Medical Center (KUMC).

**Measurements and Main Results:** On our internal hold-out set (n=53,048), the full XGBoost model reached a ROC-AUC of 0.791 for any OSA and 0.768 for moderate-severe OSA. The minimal model retained most performance (0.764 and 0.736). Temporal performance was stable. Externally, frozen models showed lower AUCs (0.718 and 0.690 for full models; 0.725 and 0.706 for minimal models) but similar to KUMC-native models and improved further with transfer learning. The models significantly outperformed STOP-BANG (AUC difference 0.06-0.09 for predicting any OSA; p<0.001). Bayesian updating with select patient-reported items further improved discrimination.

**Conclusions:** Machine learning models based on EHR data can predict OSA probability that outperform existing screeners, transfers across systems, and offers potential for EHR implementation.

**At a Glance Commentary:** *Scientific Knowledge on the Subject:* Obstructive sleep apnea is common and heavily underdiagnosed, yet upstream patient identification still depends on symptom and anthropometry questionnaires such as STOP-BANG, which are effort-dependent and achieve modest performance. Prior machine learning models for OSA are heterogeneous, often developed on small or demographically narrow cohorts, frequently rely on physiologic signals that reintroduce a testing step, and are rarely validated across independent health systems.

*What This Study Adds to the Field:* We developed machine learning models from routinely collected electronic health record data, comprising a full (106-feature) and a minimal (4-feature) model, to predict any OSA and moderate-severe OSA. The study is distinguished by multiple validation efforts, including temporal validation and external validation at a second health system. We further explored adaptation strategies for external adoption using transfer learning techniques. Finally, the models outperformed existing screeners such as STOP-BANG, and we describe a Bayesian framework that optionally incorporates select patient-reported symptoms to refine individual risk estimates. Overall, these models are feasible to implement within EHR systems as a low-effort, passive screener that can support encounter-based and population health outreach screening strategies.

## Introduction

Obstructive sleep apnea (OSA) is a public health condition affecting approximately 1 billion adults worldwide(1). OSA carries significant consequences including daytime impairment with increased accident risk, cardiometabolic disease, cognitive impairment, and substantial healthcare costs(2–6). Despite its high prevalence and consequences, OSA remains under-diagnosed, with an estimated 80% of affected individuals unrecognized(7). This disparity is driven by a lack of systematic or standardized screening and difficulty recognizing those at risk due to heterogeneous OSA phenotypes and presentations(8–11).

Questionnaire-based tools (e.g. STOP-BANG) require active patient and clinician effort, are subject to survey fatigue, and demonstrate variable performance with limited severity stratification(12–14). Artificial intelligence (AI) approaches, such as machine learning (ML), have emerged and often outperform traditional screening tools(15). However, they commonly rely on resource-intensive features for data collection (e.g. questionnaires, physical examination, and physiologic signals) which are impractical for large-scale deployment. Further, many models were trained on relatively small cohorts with limited demographic diversity and unproven generalizability(16). The lack of robust external validation was a key limitation across most studies.

OSA predictions based solely on routinely collected electronic health record (EHR) data have the potential to be minimal-effort screening tools. They can be deployed within EHR systems and alert clinicians when an at-risk patient is identified during a clinic encounter or as a population screening strategy. In this study, we developed ML models to predict OSA using typically available EHR data and validated them in an external clinical cohort. We also evaluated transfer learning approaches to provide guidance on adaptation strategies for deployment in other health systems. Finally, we described implementation strategies including a sequential risk-updating framework utilizing patient-reported data.

## Methods

### Study design

This is an observational retrospective longitudinal multi-site study. We developed and validated models for identifying OSA risk from routinely-collected EHR data (Epic Systems, Verona, Wisconsin, at both study sites), following the TRIPOD+AI guideline for machine learning prediction models(17). The study was conducted under protocols approved by the BIDMC Institutional Review Board (#2024P000804/#2022P000417), which granted a waiver of informed consent for this retrospective study. The work comprised three phases (Figure 1): model development with internal and temporal validation at the development health system (Phase 1); external validation and comparison of transfer learning strategies at an independent system (Phase 2); and comparison against existing clinical screeners and applied a sequential Bayesian risk-updating framework (Phase 3).

**Figure 1-.**
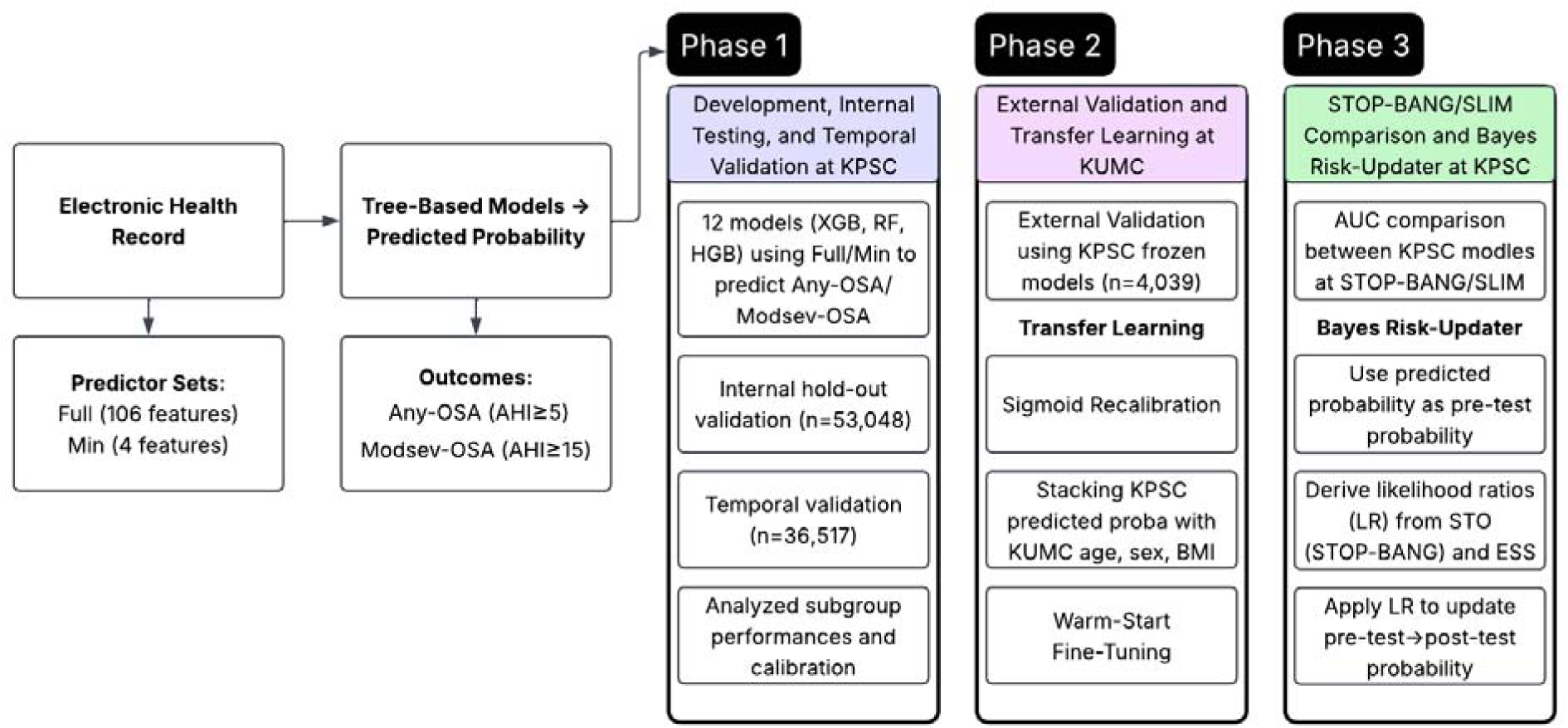
Study Design. This study comprises three phases: 1) model development, internal testing, and temporal validation; 2) external validation and transfer learning adaptation; and 3) performance comparison against current screeners and implementation considerations using a risk-updating Bayesian framework utilizing patient-reported data.

### Phase 1: Development, internal testing, and temporal validation

#### Data source and participants

Adults (age≥18 years at sleep study) who underwent a diagnostic home sleep apnea test (HSAT) or polysomnography (PSG) between February 11, 2016 and May 17, 2025 at Kaiser Permanente Southern California (KPSC) were included for analysis. Index date was defined as the sleep study date (earliest date used if multiple studies were performed). The *development* cohort was partitioned before any modeling by stratified random sampling into 80%/20% training and internal validation sets. A same-system *temporal* cohort comprised patients studied later in time, between May 18, 2025 and February 17, 2026 (duplicate patients were removed). The external validation cohort was composed of adults who underwent a diagnostic HSAT or PSG between July 23, 2005 and April 30, 2025 at the Sleep Disorders Center, University of Kansas Medical Center (KUMC).

#### Outcome

The reference standard was the apnea-hypopnea index using the 4% hypopnea definition (AHI4%). Two binary outcomes were derived: any OSA (AHI≥5, denoted as Any-OSA) and moderate-severe OSA (AHI≥15, denoted as Modsev-OSA).

#### Predictors and preprocessing

Two predictor sets were defined. The *Full* set comprised 106 EHR features spanning four domains: (i) demographics (including age, sex, and race/ethnicity); (ii) vital signs (including body-mass index [BMI]); (iii) laboratory values (39 metrics); (iv) comorbidities based on derived Charlson and Elixhauser conditions and their summary indices(18, 19). This is a validated approach for identifying comorbidities rather than utilizing unreliable EHR problem lists(20). Individual Charlson and Elixhauser conditions were identified based on International Classification of Disease (ICD9/10) coding of all available encounters for each patient; Elixhauser conditions that were redundant with Charlson were removed resulting in a unified comorbidity list. In contrast, the *Minimal* set comprised 4 feature types: age, sex, BMI, and race/ethnicity. Because missingness in EHR data may be informative, values were left unimputed and handled natively by the tree-based models. Data dictionary and cleaning parameters are included in Supplement S1.

#### Preliminary exploratory data analysis (EDA)

Prior to formal model development, we conducted EDA on a subset of the development cohort (n=26,524), which did not overlap with any other validation cohorts (Supplement S2). The base model incorporated the feature categories described above: demographics, vital signs (including BMI), comorbidities, and laboratory values. We evaluated several modeling combinations: applying probabilistic principal component analysis (PPCA) for dimensionality reduction, excluding patients with mild OSA (AHI 5-14.9) to optimize the training signal, incorporating medications as features (labeled by the first two digits of their Generic Product Identifier codes), and removing comorbidities or laboratory values from training. Based on these exploratory analyses, we adopted the base model as our Full model, did not apply PPCA, did not include medications as a feature type, and retained patients with mild OSA in the training dataset.

#### Model development and tuning

We evaluated three tree-ensemble families (selected for their inherent ability to handle missingness and their amenability to implementation): extreme gradient boosting (XGBoost [XGB]); histogram-based gradient boosting (HGB) with positive monotonic constraints for age and BMI; and Random Forests (RF). Each family was trained for both outcomes (Any-OSA, Modsev-OSA) with both predictor sets (Full and Minimal), yielding 12 models. Hyperparameters were tuned and discrimination estimated by nested cross-validation: a 5-fold stratified outer loop, with an inner 3-fold stratified loop for tuning. Bayesian optimization (50 iterations per inner search) was used to optimize the area under the receiver-operating-characteristic curve (ROC-AUC); search spaces and optimal parameters are provided in Supplement S3. For each model, the best configurations from the five outer folds were aggregated by taking the median of each hyperparameter, and the final estimator was refit on the full training set.

#### Evaluation framework

The primary performance metric was ROC-AUC, computed on out-of-fold predictions pooled across the outer folds internally and on held-out predictions at each validation set, with 95% confidence intervals (CIs) obtained by the DeLong method for correlated areas. Calibration was assessed by the calibration slope, intercept, calibration-in-the-large, and expected calibration error over ten equal-frequency bins, with reliability diagrams using Wilson CIs. Threshold metrics (sensitivity, specificity, positive/negative predictive values) were reported by fixing sensitivity to 80% and at a per-model operating threshold obtained by maximizing Youden’s J statistic on the development held-out test set; these thresholds were then applied to the external validation cohort. Clinical utility was assessed by decision-curve analysis (DCA)(21, 22). Subgroup AUCs were stratified by race/ethnicity, sex, age (≥65 vs. <65 years), and study modality (HSAT vs. PSG). Feature importance was quantified using Shapley values on the development training cohort.

#### Temporal validation and recalibration

The 12 trained (i.e., frozen) models were applied to the temporal cohort and their discrimination and calibration compared with development estimates. For recalibration, sigmoid scaling was utilized for all models with pre-specified training-validation split. This procedure was reused in Phase 2.

### Phase 2: External validation and transfer learning (KUMC)

#### External validation of the frozen models

XGB models were selected for external validation at KUMC based on performance and their amenability for warm-start fine-tuning (WSFT) transfer learning. The external cohort was partitioned by a stratified 80%/20% split. All KPSC model performances were evaluated on the KUMC test set for fair comparison against their KUMC-natively trained model counterparts. Performance was summarized using the evaluation framework described in Phase 1.

#### Transfer learning for external adaptation

Three adaptation strategies were compared at KUMC against the no-adaptation frozen models, all using the same stratified 80%/20% split. First, sigmoid recalibration was applied to KUMC predicted probabilities. Second, WSFT extended the four frozen boosters through continued training(23). WSFT is a method enabling continued training of a frozen model without merging datasets (Supplement S11). Across 10 cumulative rounds, round *k* was trained on a random split *k* × 10% of the external training set and added 20 trees per round, for a total of 200. After each round, performance was re-evaluated on the test set. The fine-tuning learning rate was set to the source rate scaled by a multiplier and clamped to [0.001, 0.05]. The KPSC model thresholds (derived from Youden’s J statistic and a fixed 80% sensitivity) were applied, and Youden’s J was also recalculated for KUMC on the training cohort. Third, we trained an XGB meta-model by stacking the KPSC frozen model predicted probability with KUMC age, sex, and BMI(24). The meta-model was refit on the full external training set before evaluation. As an additional reference, KUMC site-native XGB models were also trained using each predictor set for both outcomes applying the same nested cross-validation approach from Phase 1.

### Phase 3: Screener comparison and sequential risk-updating

#### Comparison with clinical screeners

The Full and Minimal models were compared against two existing OSA screeners on a common held-out cohort restricted to patients with full STOP-BANG data (n=1,981; baseline characteristics in Supplement S9); patients were classified as at-risk when ≥3 for Any-OSA and ≥5 for Modsev-OSA. Snore (S), tiredness (T), observed apneas (O), and neck circumference (N, collar size ≥16 vs. <16) were patient-reported; the other features were derived from EHR(25). We also compared against SLIM, a screener that utilizes EHR-derived age, sex, hypertension status, and BMI (Supplement S4)(26). Discrimination was compared using the DeLong test for correlated AUCs (paired percentile bootstrap 95% CI; 2,000 bootstraps); specificity at the STOP-BANG and SLIM sensitivity thresholds was compared using McNemar’s test. Clinical utility was compared using DCA.

#### Sequential Bayesian risk-updating

Questionnaire items were excluded from model training to preserve the zero-effort property of the EHR-only model; instead, they were evaluated in a two-stage Bayesian framework. The calibrated XGB probability served as the pre-test probability, which was then updated via Bayes’ rule using select patient-reported items and likelihood ratios (LR) estimated on the training set. Two items were used: the “STO” sub-score (the sum of the snore, tiredness, and observed-apnea items from STOP-BANG, ranging 0–3) and the Epworth Sleepiness Scale (ESS, dichotomized at >10 vs. ≤10). “P-BANG” items were excluded to avoid redundancy with existing XGB features(27). LRs were calculated with a Haldane continuity correction and Simel-method standard errors for log-LR CIs. Updated post-test probabilities were compared against pre-test probabilities by AUC (DeLong test and paired bootstrapping).

## Results

### Phase 1: Model development and internal validation (KPSC)

The KPSC development cohort comprised 265,236 adults (age 51.7±15.1 years, 56.7% male, BMI 33.7±8.1 kg/m^2^; Any-OSA prevalence 72.8%, Modsev-OSA 46.2%), partitioned into training (n=212,188) and held-out test (n=53,048) sets (both nearly identical across all characteristics), and a later same-site temporal cohort of 36,517 (Table 1).

**Table 1-.**
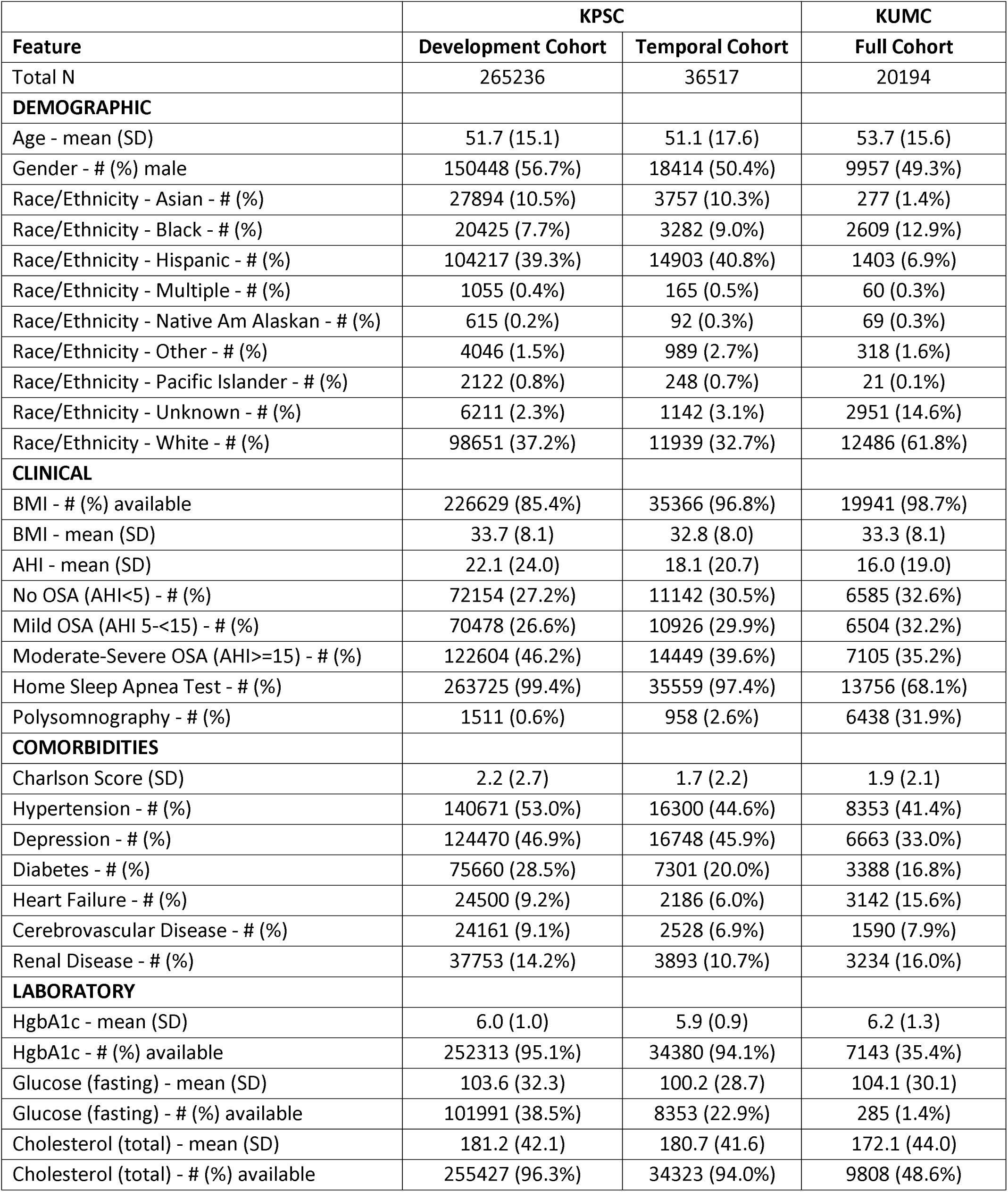
Baseline Characteristics for the Development, Temporal, and External Validation Cohorts.

|  | KPSC |  | KUMC |
| --- | --- | --- | --- |
| Feature | Development Cohort | Temporal Cohort | Full Cohort |
| Total N | 265236 | 36517 | 20194 |
| **DEMOGRAPHIC** |  |  |  |
| Age - mean (SD) | 51.7 (15.1) | 51.1 (17.6) | 53.7 (15.6) |
| Gender - # (%) male | 150448 (56.7%) | 18414 (50.4%) | 9957 (49.3%) |
| Race/Ethnicity - Asian - # (%) | 27894 (10.5%) | 3757 (10.3%) | 277 (1.4%) |
| Race/Ethnicity - Black - # (%) | 20425 (7.7%) | 3282 (9.0%) | 2609 (12.9%) |
| Race/Ethnicity - Hispanic - # (%) | 104217 (39.3%) | 14903 (40.8%) | 1403 (6.9%) |
| Race/Ethnicity - Multiple - # (%) | 1055 (0.4%) | 165 (0.5%) | 60 (0.3%) |
| Race/Ethnicity - Native Am Alaskan - # (%) | 615 (0.2%) | 92 (0.3%) | 69 (0.3%) |
| Race/Ethnicity - Other - # (%) | 4046 (1.5%) | 989 (2.7%) | 318 (1.6%) |
| Race/Ethnicity - Pacific Islander - # (%) | 2122 (0.8%) | 248 (0.7%) | 21 (0.1%) |
| Race/Ethnicity - Unknown - # (%) | 6211 (2.3%) | 1142 (3.1%) | 2951 (14.6%) |
| Race/Ethnicity - White - # (%) | 98651 (37.2%) | 11939 (32.7%) | 12486 (61.8%) |
| **CLINICAL** |  |  |  |
| BMI - # (%) available | 226629 (85.4%) | 35366 (96.8%) | 19941 (98.7%) |
| BMI - mean (SD) | 33.7 (8.1) | 32.8 (8.0) | 33.3 (8.1) |
| AHI - mean (SD) | 22.1 (24.0) | 18.1 (20.7) | 16.0 (19.0) |
| No OSA (AHI<5) - # (%) | 72154 (27.2%) | 11142 (30.5%) | 6585 (32.6%) |
| Mild OSA (AHI 5-<15) - # (%) | 70478 (26.6%) | 10926 (29.9%) | 6504 (32.2%) |
| Moderate-Severe OSA (AHI>=15) - # (%) | 122604 (46.2%) | 14449 (39.6%) | 7105 (35.2%) |
| Home Sleep Apnea Test - # (%) | 263725 (99.4%) | 35559 (97.4%) | 13756 (68.1%) |
| Polysomnography - # (%) | 1511 (0.6%) | 958 (2.6%) | 6438 (31.9%) |
| **COMORBIDITIES** |  |  |  |
| Charlson Score (SD) | 2.2 (2.7) | 1.7 (2.2) | 1.9 (2.1) |
| Hypertension - # (%) | 140671 (53.0%) | 16300 (44.6%) | 8353 (41.4%) |
| Depression - # (%) | 124470 (46.9%) | 16748 (45.9%) | 6663 (33.0%) |
| Diabetes - # (%) | 75660 (28.5%) | 7301 (20.0%) | 3388 (16.8%) |
| Heart Failure - # (%) | 24500 (9.2%) | 2186 (6.0%) | 3142 (15.6%) |
| Cerebrovascular Disease - # (%) | 24161 (9.1%) | 2528 (6.9%) | 1590 (7.9%) |
| Renal Disease - # (%) | 37753 (14.2%) | 3893 (10.7%) | 3234 (16.0%) |
| **LABORATORY** |  |  |  |
| HgbA1c - mean (SD) | 6.0 (1.0) | 5.9 (0.9) | 6.2 (1.3) |
| HgbA1c - # (%) available | 252313 (95.1%) | 34380 (94.1%) | 7143 (35.4%) |
| Glucose (fasting) - mean (SD) | 103.6 (32.3) | 100.2 (28.7) | 104.1 (30.1) |
| Glucose (fasting) - # (%) available | 101991 (38.5%) | 8353 (22.9%) | 285 (1.4%) |
| Cholesterol (total) - mean (SD) | 181.2 (42.1) | 180.7 (41.6) | 172.1 (44.0) |
| Cholesterol (total) - # (%) available | 255427 (96.3%) | 34323 (94.0%) | 9808 (48.6%) |

In nested cross-validation on pooled out-of-fold predictions (Supplement S5), AUCs for Any-OSA/Modsev-OSA were higher for XGB (0.791/0.768) and HGB (0.790/0.766) than for RF (0.781/0.753). The Minimal models followed the same pattern but performed slightly worse overall: XGB (0.766/0.737), HGB (0.765/0.736), and RF (0.763/0.735). Subgroup AUCs were consistent across all three model families (Figure 2A). For the Full XGB model specifically, subgroup performance (Any-OSA/Modsev-OSA) was 0.781–0.797/0.758–0.773 across race/ethnicity, 0.773–0.783/0.746–0.750 by sex, and 0.773–0.792/0.767–0.783 by study modality (HSAT vs PSG). Discrimination, however, was lower in patients ≥65 versus <65 years (0.673 vs. 0.810 for Any-OSA and 0.673 vs. 0.788 for Modsev-OSA). The decrease in performance with increasing age was gradual (Figure 2B). Subgroup patterns were consistent across all models and outcomes.

**Figure 2-.**
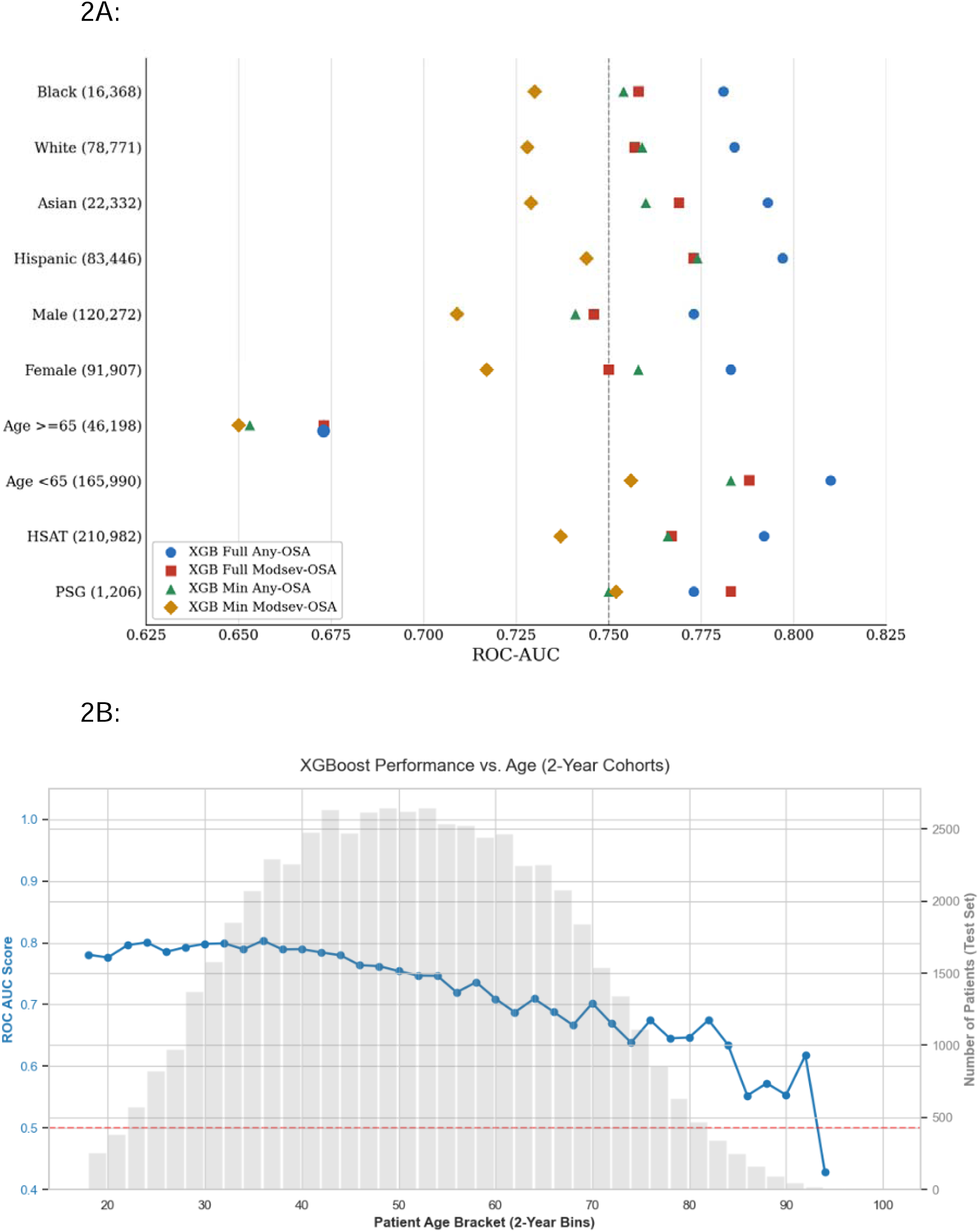
XGBoost Subgroup Performances. The XGBoost models demonstrated consistent performance across race/ethnicity, sex, and study modality subgroups, with the exception being lower performance among participants aged ≥65 years (Figure 2A). The decrease in performance with increasing age was gradual (Figure 2B).

On the held-out test set (Table 2), performances across all families, predictor sets, and outcomes were preserved with slightly higher AUC values than the cross-validation results. On the temporal cohort, discrimination was again preserved. XGB and HGB continued to outperform RF. Although models were already well calibrated to the temporal cohort, sigmoid recalibration was applied to all 12 frozen models (Supplement S6). XGB was selected as the primary model due to performance and its greater amenability than HGB for transfer learning. Classification performance metrics are reported for Youden’s J-determined optimal threshold and when sensitivity is fixed to 80% in Supplement S10. The optimal threshold revealed a balanced sensitivity (around 75%) and specificity (around 65%) for both Any-OSA and Modsev-OSA, although metrics for the Minimal Modsev-OSA model were slightly worse. Across both OSA outcomes (Any-OSA and Modsev-OSA), age, BMI, and sex were the top three predictors in both XGB models based on Shapley values, comprising approximately 35% and 88% of total feature attribution in the Full and Minimal models, respectively (Supplement S7). Of the Full models’ four feature categories (demographics, labs, vitals, and comorbidities), labs were the most important for both Any-OSA and Modsev-OSA (37.6% and 36.6% of overall feature attribution), followed by demographics (27.9% and 27.8%), vitals including BMI (23.1% and 24.3%), and comorbidities (11.2% and 11.2%).

**Table 2-.** Performance On Internal Hold-out Set and Temporal Validation. Performance across all models was sustained in both the internal hold-out set and temporal validation sets with almost identical AUCs between the two. The Full predictor set comprises 106 features across demographics, labs, comorbidities, and vitals (including BMI). The Minimal predictor set comprises 4 features including age, sex, BMI, and race/ethnicity.

| Model | Target | Hold Out | Temporal |
| --- | --- | --- | --- |
| Full |  | AUC (95% CI) | AUC (95% CI) |
| XGBoost | Any OSA | 0.790 (0.786–0.794) | 0.788 (0.783–0.793) |
| RF |  | 0.773 (0.768–0.777) | 0.774 (0.769–0.779) |
| HGB |  | 0.789 (0.784–0.793) | 0.788 (0.783–0.793) |
| XGBoost | Modsev OSA | 0.768 (0.764–0.772) | 0.768 (0.763–0.772) |
| RF |  | 0.750 (0.746–0.754) | 0.751 (0.746–0.756) |
| HGB |  | 0.766 (0.762–0.770) | 0.767 (0.762–0.772) |
| Minimal |  |  |  |
| XGBoost | Any OSA | 0.764 (0.760–0.769) | 0.770 (0.765–0.775) |
| RF |  | 0.761 (0.756–0.766) | 0.769 (0.763–0.774) |
| HGB |  | 0.763 (0.759–0.768) | 0.770 (0.765–0.775) |
| XGBoost | Modsev OSA | 0.736 (0.732–0.740) | 0.742 (0.737–0.748) |
| RF |  | 0.733 (0.729–0.738) | 0.742 (0.737–0.747) |
| HGB |  | 0.735 (0.731–0.740) | 0.743 (0.738–0.748) |

### Phase 2: External validation and transfer learning (KUMC)

The four frozen XGB models were transferred to the KUMC cohort (n=20,194; train 16,155/test 4,039). Compared to KPSC, this cohort was slightly older (age 53.7±15.6), more White (61.8%) with fewer Hispanic (6.9%) and Asian (1.4%) patients, had lower OSA burden (AHI4%≥5 67.4%, AHI4%≥15 35.2%), and sparser laboratory coverage (HbA1c available 35.4% vs 95.1%) (Table 1). The KUMC training and test sets had nearly identical baseline characteristics.

Initial testing of the frozen models yielded performance comparable to, though slightly lower than, the natively trained KUMC XGBoost models (Table 3), demonstrating the generalizability of the KPSC models. Minimal models surprisingly slightly outperformed Full models (Any-OSA [0.725 vs. 0.718]; Modsev-OSA [0.706 vs. 0.690]). We also performed a sensitivity analysis that compared performance on a cohort of patients (n=7,998) that had both AHI3% and AHI4% derived from the same polysomnography study, which revealed similar results for both metrics (Supplement S9); for example, AUC for AHI3% was actually higher than for AHI4% for predicting Any-OSA (Full model 0.747 vs. 0.735; Min model 0.758 vs. 0.733). Although models were already well calibrated to the frozen cohort, sigmoid recalibration was applied to all 4 frozen models (Supplement S6). Classification performance metrics after model recalibration at the KPSC Youden’s J-determined optimal threshold and when fixed to 80% sensitivity revealed decreased sensitivities and increased specificities for both Any-OSA and Modsev-OSA (Table 3). Re-determining optimal threshold at KUMC rebalanced sensitivities and specificities.

**Table 3-.**
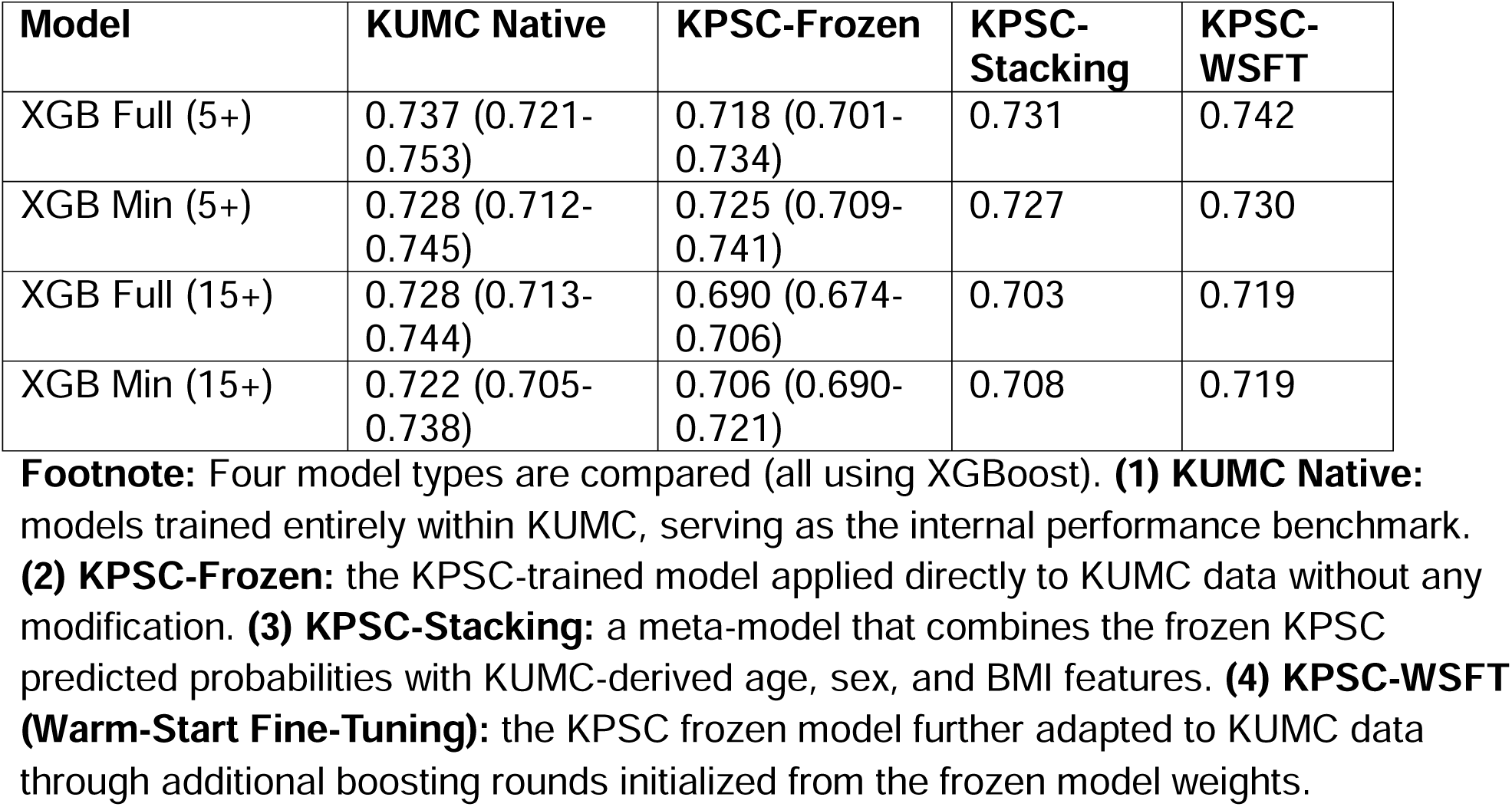
External Validation and Transfer Learning. When KPSC frozen models were initially tested on KUMC, performance was slightly lower than but comparable to KUMC natively trained models. We evaluated two adaptation techniques: warm-start fine-tuning improved performance more than stacking, particularly for the Full models.

| Model | KUMC Native | KPSC-Frozen | KPSC-Stacking | KPSC-WSFT |
| --- | --- | --- | --- | --- |
| XGB Full (5+) | 0.737 (0.721-0.753) | 0.718 (0.701-0.734) | 0.731 | 0.742 |
| XGB Min (5+) | 0.728 (0.712-0.745) | 0.725 (0.709-0.741) | 0.727 | 0.730 |
| XGB Full (15+) | 0.728 (0.713-0.744) | 0.690 (0.674-0.706) | 0.703 | 0.719 |
| XGB Min (15+) | 0.722 (0.705-0.738) | 0.706 (0.690-0.721) | 0.708 | 0.719 |

**Footnote:** Four model types are compared (all using XGBoost). **(1) KUMC Native:** models trained entirely within KUMC, serving as the internal performance benchmark. **(2) KPSC-Frozen:** the KPSC-trained model applied directly to KUMC data without any modification. **(3) KPSC-Stacking:** a meta-model that combines the frozen KPSC predicted probabilities with KUMC-derived age, sex, and BMI features. **(4) KPSC-WSFT (Warm-Start Fine-Tuning):** the KPSC frozen model further adapted to KUMC data through additional boosting rounds initialized from the frozen model weights.

Stacking and WSFT transfer learning methods both demonstrated improved performance (Table 4). Initially, for several of the models, the performance dropped (Figure 3). But with incremental addition of each 10% tranche (n=1616) of patients for training, performance gradually improved with all models eventually exceeding their corresponding KPSC frozen model performance. However, both the stacking approach and Minimal models plateaued around 60% (∼10,000 patients), while both Full WSFT models (Any-OSA and Modsev-OSA) continued to increase at a linear rate with no signs of plateauing. For Any-OSA, the Full WSFT and both Minimal models outperformed their KUMC native counterparts.

**Figure 3-.**
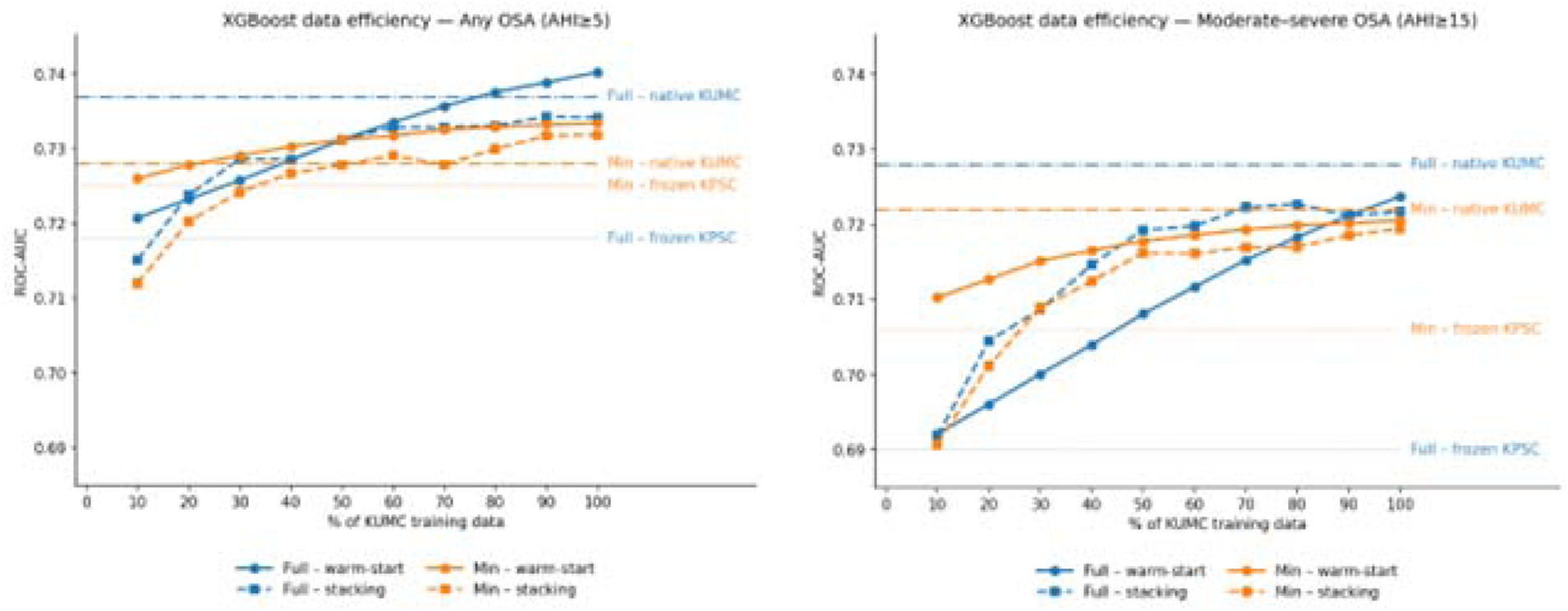
Transfer Learning Data Efficiency Analysis Comparing Stacking and Warm-Start Fine-Tuning. Frozen KPSC models underwent transfer learning on the KUMC training set (n=16,155), with data efficiency assessed by measuring performance at each successive 10% increment (n=1,616) and AUCs compared against the frozen KPSC and KUMC-native models. Full (blue) and Minimal (orange) models were fine-tuned via warm-start (solid) and stacking (dashed); horizontal lines mark the native KUMC and frozen KPSC benchmarks. All transfer learning models eventually surpassed their frozen KPSC counterpart. Stacking and Minimal models plateaued near 60% of training data (∼10,000 patients), while both Full warm-start models showed near-linear improvement without plateauing. For Any-OSA, three of the models outperformed their native KUMC counterparts.

### Phase 3: Comparison with clinical screeners and Bayesian risk-updating

On the KPSC common held-out cohort with complete STOP-BANG data (n=1,981; baseline characteristics in Supplement S8), the XGB models out-performed both STOP-BANG and SLIM on both outcomes (Figure 4). AUC performance across XGB Full/Minimal, STOP-BANG, and SLIM are as follows: for Any-OSA (0.771/0.750 vs. 0.690 and 0.662) and for Modsev-OSA (0.743/0.724 vs. 0.706 and 0.645); all comparisons except XGB Minimal Modsev-OSA versus STOP-BANG (p=0.099) were statistically significant.

**Figure 4-.**
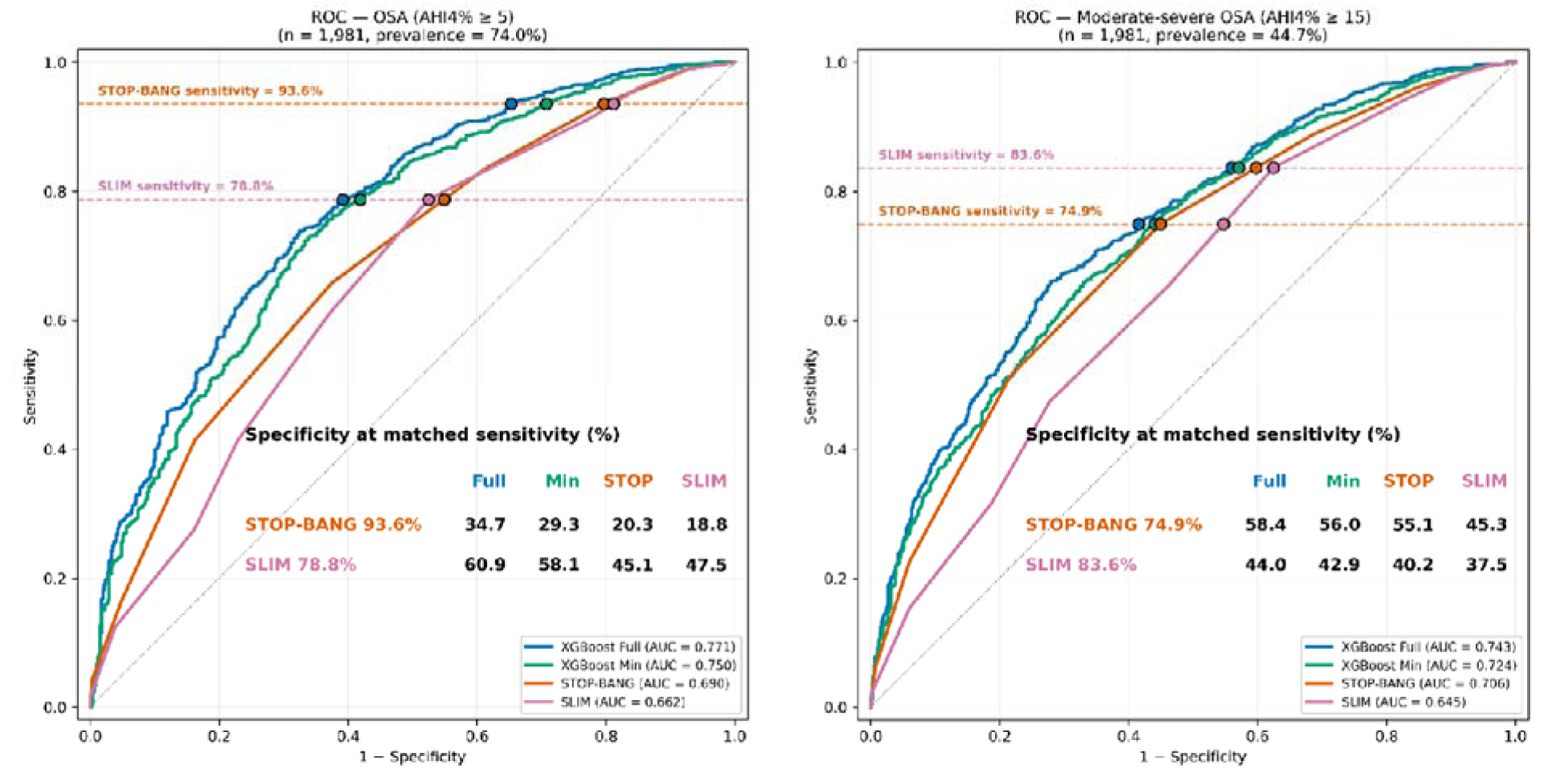
Comparison of ROCs and Specificities Across Screeners. Both Full and Minimal XGBoost models had better AUCs than STOP-BANG and SLIM. Using thresholds for STOP-BANG (≥ 3 for Any OSA and ≥ 5 for Mod-severe OSA) and SLIM (> 1 for both outcomes), both XGBoost models demonstrated superior specificities at matched sensitivities.

When matching XGB sensitivity to both screener’s sensitivity, specificity was consistently higher for XGB models; all specificity comparisons were statistically significant (McNemar p<0.001), except for comparing XGB Min Modsev-OSA to STOP-BANG (p=0.06) (Figure 4). On decision-curve analysis, the ML models demonstrated higher net benefit across all thresholds (Figure 5). Further, the XGB models’ net benefit was higher than Treat All across the entire spectrum of thresholds, supporting its clinical utility.

**Figure 5-.**
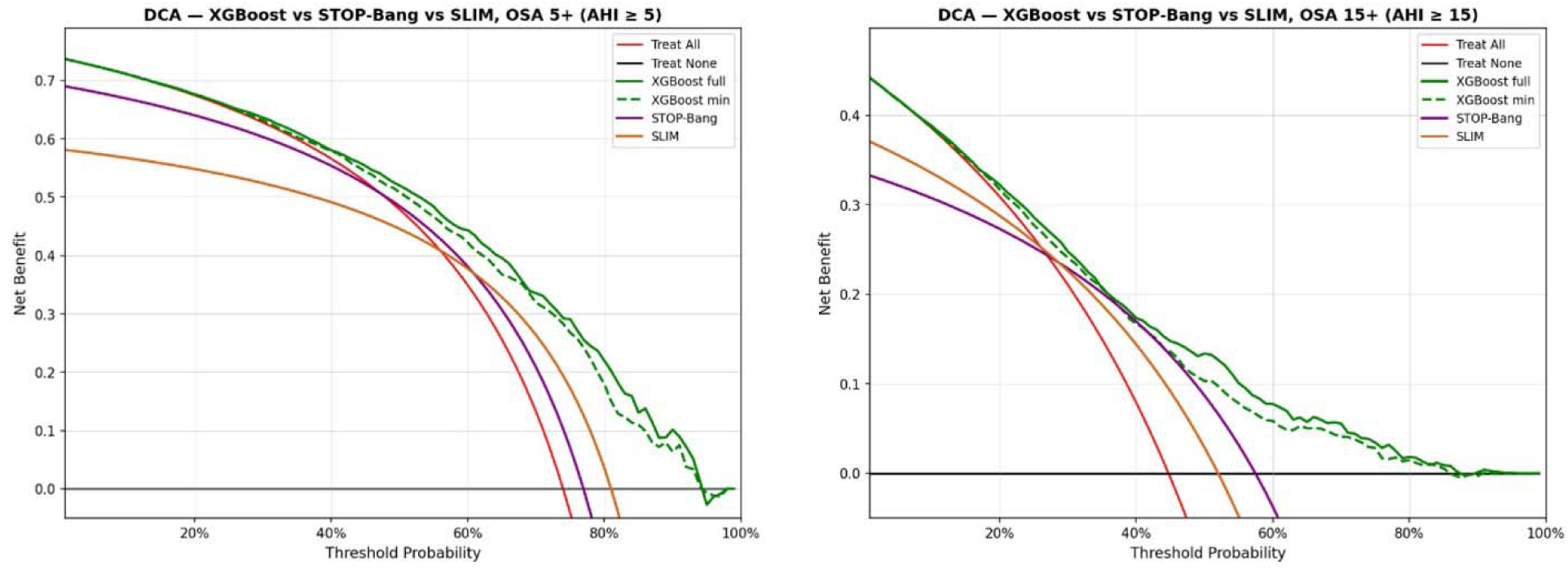
Decision Curve Analysis to Compare Clinical Utility Between Screeners. Decision Curve Analysis (DCA) is a plot of Net Benefit (the proportion of true positives minus the weighted cost of false positives) for a given threshold. Both Full and Minimal models shows consistently better net benefit than STOP-BANG, SLIM, and the “Treat All” curve across all thresholds.

Sequential Bayesian updating of calibrated XGB probabilities was evaluated on a held-out cohort (n=1981, baseline characteristics in Supplement S8). STO and ESS likelihood ratios for Any-OSA/Modsev-OSA were: STO=0 or 1 (06.2/0.56); STO=2 (0.97/0.93); STO=3 (1.89/1.85); ESS>10 (1.20/1.27); ESS≤10 (0.96/0.93). STO significantly improved AUC for all four models. ESS updating also revealed statistically significant pre-post gains in AUC, however, the pre-post difference was negligible and likely not meaningful for implementation (Figure 6). All pre-post comparisons were statistically significant (p<0.05).

**Figure 6-.**
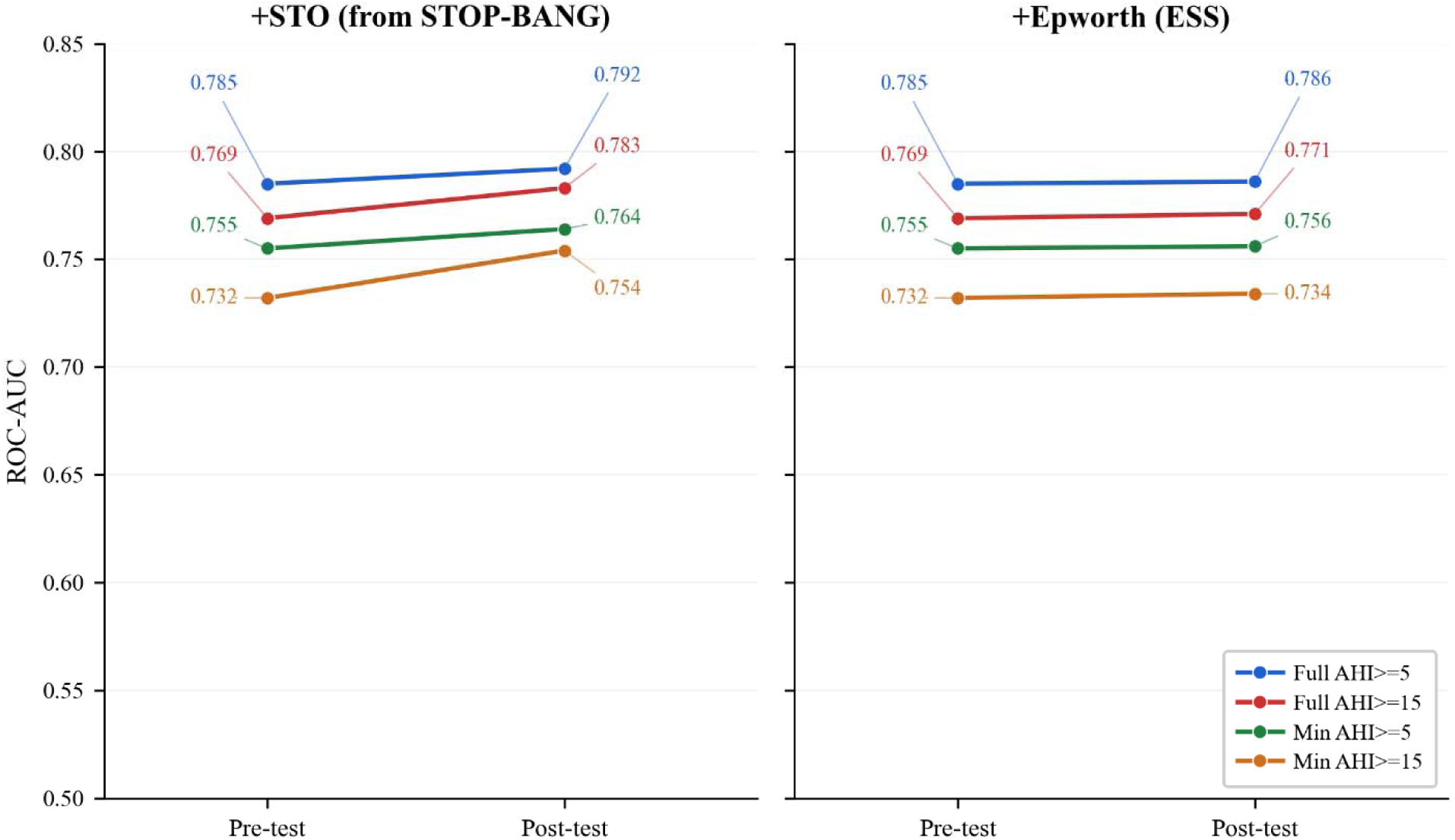
Bayesian Risk-Updating Framework using Patient-Reported Data to Improve Model Performance. Likelihood ratios were derived from “STO” elements from STOP-BANG (score 0-3) and Epworth Sleepiness Scale (>10 vs. ≤10) to update the model’s predicted probability. Post-test model performance improved to a greater degree through STO than ESS (which was negligible). All pre-post AUC comparisons were statistically significant (p<0.05).

## Discussion

The aim of this study was to develop ML prediction models utilizing EHR data as screening tools for any OSA and moderate-to-severe OSA. Our key findings are as follows. First, the models discriminated at-risk patients with sufficient performance to serve as screening or triage tools, with performance validated across patient subgroups. Second, minimal models using only age, sex, BMI, and race/ethnicity retained nearly full performance, offering an option that may be easier to implement. Third, the models generalized to an external health system, and were further improved with transfer learning. Fourth, the models outperformed STOP-BANG, the most common existing OSA screener(25). Adding select patient-reported symptoms through a sequential Bayesian risk-updating framework improved performance further.

These findings reflect several conceptual and methodological strengths. The development cohort was large (265,236 patients) and demographically diverse, which both stabilizes estimation and allowed discrimination to be examined directly across race/ethnicity, sex, age, and study modality. External validation in another clinical setting, typically not performed among previously published ML-based OSA studies, was conducted at a site with a different demographic mix, comorbidity profile, and laboratory feature availability, supporting generalizability to heterogeneous populations and clinical practices(15). The models were also designed to use pre-existing EHR data (without requiring additional data collection such as questionnaires, physical examination, or physiologic signals [e.g., electrocardiogram]), making them a zero-effort screening tool requiring no additional patient or clinician effort. Compared with a similar artificial neural network (ANN)-based model that also used EHR-only data, our models demonstrated better performance (the ANN [AUC 0.68] did not outperform STOP-BANG [AUC 0.71])(28). Further, the ANN also incorporated fewer overall and less diverse race/ethnicity features, conceptually offers less interpretability, handles missingness inherent in real-world EHR data with less flexibility, and may be more complex to deploy.

Several characteristics of our prediction models support clinical implementation. The algorithm is designed to run passively within the EHR and can be deployed in multiple ways. It can support encounter-based screening, such as surfacing risk during a primary-care visit, or population-based screening in which the algorithm is applied to a patient panel followed by dedicated outreach. To make the prediction output actionable, a risk probability or flag could appear within a patient’s EHR profile. An effective screening strategy, however, requires an active patient engagement component beyond merely patient risk identification. Our previous experience with primary care-based sleep disorder screening revealed that most patients identified to be at-risk failed to keep their sleep center appointment after referral(29). Finally, while questionnaire items were deliberately excluded from the models to preserve their zero-effort property, we incorporated patient-reported “STO” symptoms to update risk probability through a Bayesian framework. The number of positive “STO” symptoms can update the model-derived probability through a calculator that applies their likelihood ratios; alternatively, a clinician can crudely re-estimate risk by mentally doubling the post-test odds when a patient reports all three symptoms.

Adoption of the prediction models at external systems involves several considerations. First, the study demonstrated generalizability of KPSC models; while AUC values were lower than when tested internally at KPSC, they were comparable to those of KUMC-natively trained models. Therefore, the KPSC frozen models can reasonably be implemented without additional training. Second, the Minimal models unexpectedly outperformed the Full models at the external site, suggesting that the three demographic features and BMI transported more reliably than the sparse, site-specific laboratory and comorbidity features. The Minimal model may therefore be preferred for external adoption, with the added advantage of simpler implementation. Third, transfer learning improved performance across all models, with most (for predicting Any-OSA) surpassing their counterparts trained natively at KUMC. The greatest improvement came from WSFT applied to the full-feature dataset, where performance had not yet plateaued, suggesting that additional training data could have produced further gains. Fourth, recalibration and re-determining thresholds to optimize sensitivity/specificity may be necessary when implementing.

This study has limitations. First, the models were developed in patients who had an indication for a sleep study and are therefore not representative of a general or primary-care population. Recalibration may be necessary before broad implementation; while outside the scope of this study, potential approaches include drawing on prospective sleep testing in the general population, calibrating to the prevalence of the target population using existing epidemiologic literature(30–34), or other posterior probability adjustment algorithms(35). Another limitation involves performance degradation with increasing age, which we suspect reflects discriminating features (e.g. hypertension) that are informative in younger patients may become less discriminatory in older patients due to general increase in disease burden. Furthermore, models were trained to detect AHI4% and not other hypopnea definitions, although our sensitivity analysis on a population at KUMC with both AHI4% and AHI3% revealed similar performance. Finally, the study is retrospective and requires prospective validation.

In conclusion, machine learning models based on EHR data can identify patients at risk of any OSA and moderate-to-severe OSA. These models represent minimal-effort algorithms designed for feasible implementation in EHR systems and adoption across different health systems. Prospective validation of their implementation is required.

## Supporting information

All Supplementals

## Data Availability

The individual-level data underlying this study are not publicly available because they contain protected health information. All aggregate results supporting the findings of this study are provided within the manuscript and its supplementary materials.

## Author Contributions

Nathanael H. Hwang: All aspects of study

Diego R. Mazzotti: All aspects of study

Soniya Mishra: All aspects of study

M. Brandon Westover: All aspects of study

Dennis Hwang: All aspects of study

Robert J. Thomas: Analysis, Interpretation, Reviewing

Emmanuel Mignot: Analysis, Interpretation, Reviewing

Umakanth Katwa: Analysis, Interpretation, Reviewing

Jiaxiao Shi: Acquisition, Reviewing

Rui Yan: Acquisition, Reviewing

Kendra A. Becker: Acquisition, Reviewing

Matthew T. Klimper: Acquisition, Reviewing

Jessica K. Jara: Acquisition, Reviewing

Damien R. Stevens: Acquisition, Reviewing

Matthew K.P. Gratton: Acquisition, Reviewing

Haoqi Sun: All aspects of study

## Funding

National Institutes of Health; National Heart, Lung, and Blood Institute (NHLBI),

Bethesda, MD (R01 HL161253-01A1, P01HL160471, R01HL170675)

American Heart Association, Dallas, TX (20CDA35310360), NCATS CTSA Award UL1TR002366.

## Acknowledgments

Artificial intelligence was used to improve language and grammar and was carefully reviewed by authors for accuracy.

