## Supplementary material for "Machine Learning Models Using Electronic Health Record Data to Predict Obstructive Sleep Apnea": All Supplementals

**Supplement S1 – Data Dictionary and Processing Instructions.** This table summarizes the data structure used for the KPSC internal development site and the KUMC external validation site. It includes the data dictionary, value limits, units, and time points or censoring windows relative to the index date. Medication features (not shown) were evaluated only in exploratory models using the first two digits of the Generic Product Identifier (GPI) code, yielding 90 medication-class variables; they were excluded from final Full models because they added many predictors without meaningful performance improvement.

| FEATURE | DATA DICTIONARY | CATEGORY | Range-Min | Range-Max | Units | Value Selection |
|---|---|---|---|---|---|---|
| GENDER | Sex | Demographics | M and F | M and F | | |
| RACE_ETH | Race/ethnicity (hisp, white non-hisp, black non-hisp, Asian, Other/Multiple) | Demographics | | | | |
| age | Age | Demographics | 18 | 105 | | At time of index date |
| AHI | Apnea-Hypopnea Index | Sleep Diagnosis | 0 | 200 | per hour | Earliest if multiple |
| OSA_bin | OSA Yes/No AHI 5+ vs <5 (1/0) | Sleep Diagnosis | | | | Earliest if multiple |
| OSA_modsev | Mod-Sev OSA Yes/No AHI 15+ vs <15 (1/) | Sleep Diagnosis | | | | Earliest if multiple |
| icdx_e_ch_mi | Myocardial infarction; Charlson | Co-morbidities | 0 | 1 | | Calculated at index date |
| icdx_e_ch_chf | Congestive heart failure; Charlson | Co-morbidities | 0 | 1 | | Calculated at index date |
| icdx_e_ch_pvd | Peripheral vascular disease; Charlson | Co-morbidities | 0 | 1 | | Calculated at index date |
| icdx_e_ch_cevd | Cerebrovascular disease; Charlson | Co-morbidities | 0 | 1 | | Calculated at index date |
| icdx_e_ch_dem | Dementia; Charlson | Co-morbidities | 0 | 1 | | Calculated at index date |
| icdx_e_ch_copd | Chronic obstructive pulmonary disease; Charlson | Co-morbidities | 0 | 1 | | Calculated at index date |
| icdx_e_ch_rheum | Rheumatologic disease; Charlson | Co-morbidities | 0 | 1 | | Calculated at index date |
| icdx_e_ch_pud | Peptic ulcer disease; Charlson | Co-morbidities | 0 | 1 | | Calculated at index date |
| icdx_e_ch_mildld | Mild liver disease; Charlson | Co-morbidities | 0 | 1 | | Calculated at index date |
| icdx_e_ch_diab_nc | Diabetes without complications; Charlson | Co-morbidities | 0 | 1 | | Calculated at index date |
| icdx_e_ch_diab_c | Diabetes with complications; Charlson | Co-morbidities | 0 | 1 | | Calculated at index date |
| icdx_e_ch_para | Paralysis/hemiplegia; Charlson | Co-morbidities | 0 | 1 | | Calculated at index date |
| icdx_e_ch_rd | Renal disease; Charlson | Co-morbidities | 0 | 1 | | Calculated at index date |
| icdx_e_ch_cancer | Cancer; Charlson | Co-morbidities | 0 | 1 | | Calculated at index date |
| icdx_e_ch_msld | Moderate/severe liver disease; Charlson | Co-morbidities | 0 | 1 | | Calculated at index date |
| icdx_e_ch_mets | Metastatic solid tumor; Charlson | Co-morbidities | 0 | 1 | | Calculated at index date |
| icdx_e_ch_hiv | HIV/AIDS; Charlson | Co-morbidities | 0 | 1 | | Calculated at index date |
| Charlson_Wt | Weighted Charlson Comorbidity Index score | Co-morbidities | | | | Calculated at index date |
| icdx_e_ex_Arrhy | Arrhythmia; Elixhauser | Co-morbidities | 0 | 1 | | Calculated at index date |
| icdx_e_ex_VD | Valvular heart disease; Elixhauser | Co-morbidities | 0 | 1 | | Calculated at index date |
| icdx_e_ex_PCD | Pulmonary circulation disorder; Elixhauser | Co-morbidities | 0 | 1 | | Calculated at index date |
| icdx_e_ex_HPTN_NC | Uncomplicated hypertension; Elixhauser | Co-morbidities | 0 | 1 | | Calculated at index date |
| icdx_e_ex_HPTN_C | Complicated hypertension; Elixhauser | Co-morbidities | 0 | 1 | | Calculated at index date |
| icdx_e_ex_OthND | Other neurological disorder; Elixhauser | Co-morbidities | 0 | 1 | | Calculated at index date |
| icdx_e_ex_Hptothy | Hypothyroidism; Elixhauser | Co-morbidities | 0 | 1 | | Calculated at index date |
| icdx_e_ex_RF | Renal failure; Elixhauser | Co-morbidities | 0 | 1 | | Calculated at index date |
| icdx_e_ex_LD | Liver disease; Elixhauser | Co-morbidities | 0 | 1 | | Calculated at index date |
| icdx_e_ex_PUD_NB | Peptic ulcer disease without bleeding; Elixhauser | Co-morbidities | 0 | 1 | | Calculated at index date |
| icdx_e_ex_Lymp | Lymphoma; Elixhauser | Co-morbidities | 0 | 1 | | Calculated at index date |
| icdx_e_ex_Tumor | Solid tumor without metastasis; Elixhauser | Co-morbidities | 0 | 1 | | Calculated at index date |
| icdx_e_ex_Rheum_A | Rheumatoid/collagen vascular disease; Elixhauser | Co-morbidities | 0 | 1 | | Calculated at index date |
| icdx_e_ex_Coag | Coagulopathy; Elixhauser | Co-morbidities | 0 | 1 | | Calculated at index date |
| icdx_e_ex_Obesity | Obesity; Elixhauser | Co-morbidities | 0 | 1 | | Calculated at index date |
| icdx_e_ex_WL | Weight loss; Elixhauser | Co-morbidities | 0 | 1 | | Calculated at index date |
| icdx_e_ex_Fluid | Fluid/electrolyte disorder; Elixhauser | Co-morbidities | 0 | 1 | | Calculated at index date |
| icdx_e_ex_BLA | Blood loss anemia; Elixhauser | Co-morbidities | 0 | 1 | | Calculated at index date |
| icdx_e_ex_DA | Deficiency anemia; Elixhauser | Co-morbidities | 0 | 1 | | Calculated at index date |
| icdx_e_ex_Alcohol | Alcohol abuse; Elixhauser | Co-morbidities | 0 | 1 | | Calculated at index date |
| icdx_e_ex_Drug | Drug abuse; Elixhauser | Co-morbidities | 0 | 1 | | Calculated at index date |
| icdx_e_ex_Psycho | Psychotic disorder; Elixhauser | Co-morbidities | 0 | 1 | | Calculated at index date |

| | | | | | | |
|---|---|---|---|---|---|---|
| icdx_e_ex_Dep | Depression; Elixhauser | Co-morbidities | 0 | 1 | | Calculated at index date |
| Elixhauser | Elixhauser comorbidity index score | Co-morbidities | | | | Calculated at index date |
| BMI | Body mass index (kg/m$^2$) | Vitals | 15 | 110 | kg/m$^2$ | Closet prior to index date (Range 1 year) |
| HT | Patient height measurement. | Vitals | 48 | 85 | inches | Closet prior to index date (Range 1 year) |
| WT | Patient weight measurement. | Vitals | 50 | 800 | pounds (lb) | Closet prior to index date (Range 1 year) |
| DBP | Diastolic blood pressure (mmHg). | Vitals | 30 | 160 | mmHg | Closet prior to index date (Range 1 year) |
| SBP | Systolic blood pressure (mmHg). | Vitals | 60 | 300 | mmHg | Closet prior to index date (Range 1 year) |
| TEMPERATURE | Body temperature measurement. | Vitals | 95 | 110 | °F | Closet prior to index date (Range 1 year) |
| O2_LPM | Oxygen flow rate in liters per minute (L/min). | Vitals | 0 | 10 | L/min | Closet prior to index date (Range 1 year) |
| SPO2 | Peripheral oxygen saturation (%). | Vitals | 50 | 100 | % | Closet prior to index date (Range 1 year) |
| PULSE | Heart rate measured in beats per minute. | Vitals | 20 | 250 | bpm | Closet prior to index date (Range 1 year) |
| PEAK_FLOW | Peak expiratory flow measurement. | Vitals | 50 | 900 | L/min | Closet prior to index date (Range 1 year) |
| RESPIRATION | Respiratory rate measured in breaths per minute. | Vitals | 5 | 60 | breaths/min | Closet prior to index date (Range 1 year) |
| ALKALINE PHOSPHATASE_num | Serum alkaline phosphatase laboratory value. | Laboratory | 5 | 2000 | U/L | Closest prior to index date (Range 5 years) |
| ALT_num | Alanine aminotransferase (ALT) laboratory value. | Laboratory | 1 | 5000 | U/L | Closest prior to index date (Range 5 years) |
| AST_num | Aspartate aminotransferase (AST) laboratory value. | Laboratory | 1 | 5000 | U/L | Closest prior to index date (Range 5 years) |
| B TYPE NATRIURETIC PEPTIDE_num | B-type natriuretic peptide (BNP) laboratory value. | Laboratory | 0 | 10000 | pg/mL | Highest value prior to index date (Range 5 years) |
| BILIRUBIN, TOTAL_num | Total bilirubin laboratory value. | Laboratory | 0 | 30 | mg/dL | Closest prior to index date (Range 5 years) |
| BUN_num | Blood urea nitrogen (BUN) laboratory value. | Laboratory | 1 | 200 | mg/dL | Closest prior to index date (Range 5 years) |
| C-REACTIVE PROTEIN_num | C-reactive protein laboratory value. | Laboratory | 0 | 500 | mg/L | Closest prior to index date (Range 5 years) |
| CALCIUM_num | Serum calcium laboratory value. | Laboratory | 4 | 20 | mg/dL | Closest prior to index date (Range 5 years) |
| CHLORIDE_num | Serum chloride laboratory value. | Laboratory | 70 | 130 | mmol/L | Closest prior to index date (Range 5 years) |
| CHOLESTEROL_num | Total cholesterol laboratory value. | Laboratory | 50 | 1200 | mg/dL | Closest prior to index date (Range 5 years) |
| CHOLESTEROL, NON-HDL_num | Non-HDL cholesterol laboratory value. | Laboratory | 0 | 450 | mg/dL | Closest prior to index date (Range 5 years) |
| CHOLESTEROL/HIGH DENSITY LIPOPROTEIN_num | Total cholesterol to HDL ratio. | Laboratory | 0.5 | 20 | unitless | Closest prior to index date (Range 5 years) |
| CO2_num | Serum bicarbonate ($CO_2$) laboratory value. | Laboratory | 5 | 50 | mmol/L | Closest prior to index date (Range 5 years) |
| CREATININE_num | Serum creatinine laboratory value. | Laboratory | 0.1 | 30 | mg/dL | Closest prior to index date (Range 5 years) |
| FERRITIN_num | Serum ferritin laboratory value. | Laboratory | 1 | 10000 | ng/mL | Closest prior to index date (Range 5 years) |
| GLUCOSE, FASTING_num | Fasting blood glucose laboratory value. | Laboratory | 40 | 1100 | mg/dL | Closest prior to index date (Range 5 years) |
| GLUCOSE, RANDOM_num | Random blood glucose laboratory value. | Laboratory | 40 | 1100 | mg/dL | Closest prior to index date (Range 5 years) |
| HCT, AUTO_num | Hematocrit laboratory value. | Laboratory | 10 | 75 | % | Closest prior to index date (Range 5 years) |
| HDL_num | High-density lipoprotein cholesterol laboratory value. | Laboratory | 5 | 175 | mg/dL | Closest prior to index date (Range 5 years) |
| HGB_num | Hemoglobin laboratory value. | Laboratory | 3 | 25 | g/dL | Closest prior to index date (Range 5 years) |
| HGBA1C%_num | Hemoglobin A1c percentage laboratory value. | Laboratory | 3 | 25 | % | Closest prior to index date (Range 5 years) |
| IRON_num | Serum iron laboratory value. | Laboratory | 1 | 1000 | µg/dL | Closest prior to index date (Range 5 years) |
| IRON SAT_num | Iron saturation laboratory value. | Laboratory | 0 | 100 | % | Closest prior to index date (Range 5 years) |
| LDL_num | Low-density lipoprotein cholesterol laboratory value. | Laboratory | 0 | 500 | mg/dL | Closest prior to index date (Range 5 years) |
| MCH_num | Mean corpuscular hemoglobin laboratory value. | Laboratory | 10 | 60 | pg | Closest prior to index date (Range 5 years) |
| MCHC_num | Mean corpuscular hemoglobin concentration laboratory value. | Laboratory | 10 | 60 | g/dL | Closest prior to index date (Range 5 years) |
| MCV_num | Mean corpuscular volume laboratory value. | Laboratory | 40 | 140 | fL | Closest prior to index date (Range 5 years) |
| PLATELETS, AUTOMATED COUNT_num | Platelet count laboratory value. | Laboratory | 1 | 1000 | $\times10^3/\mu L$ | Closest prior to index date (Range 5 years) |
| POTASSIUM_num | Serum potassium laboratory value. | Laboratory | 2 | 10 | mmol/L | Closest prior to index date (Range 5 years) |
| RBC, AUTO_num | Red blood cell count laboratory value. | Laboratory | 1 | 10 | $\times10^6/\mu L$ | Closest prior to index date (Range 5 years) |
| RDW, BLOOD_num | Red cell distribution width laboratory value. | Laboratory | 8 | 50 | % | Closest prior to index date (Range 5 years) |
| SODIUM_num | Serum sodium laboratory value. | Laboratory | 110 | 180 | mmol/L | Closest prior to index date (Range 5 years) |
| TOTAL IRON BINDING CAPACITY_num | Total iron binding capacity laboratory value. | Laboratory | 50 | 1200 | µg/dL | Closest prior to index date (Range 5 years) |
| TRIGLYCERIDE_num | Triglyceride laboratory value (fasting). | Laboratory | 10 | 10000 | mg/dL | Closest prior to index date (Range 5 years) |
| TRIGLYCERIDE, NONFASTING_num | Triglyceride laboratory value (non-fasting). | Laboratory | 10 | 10000 | mg/dL | Closest prior to index date (Range 5 years) |
| TROPONIN I_num | Troponin I cardiac biomarker laboratory value. | Laboratory | 0 | 100 | ng/mL | Highest value prior to index date (Range 5 years) |
| TSH_num | Thyroid-stimulating hormone laboratory value. | Laboratory | 0.01 | 100 | µIU/mL | Closest prior to index date (Range 5 years) |
| WBC'S AUTO_num | White blood cell count laboratory value. | Laboratory | 0.1 | 250 | $\times10^3/\mu L$ | Closest prior to index date (Range 5 years) |

**Supplement S2 – Preliminary Exploratory Data Analysis (EDA).** Prior to formal model development, preliminary exploratory modeling on a subset (n=26,524) of the development cohort was performed. The base model included demographics, vitals including BMI, comorbidities, and laboratory values. We then tested different combinations of modeling approaches: 1) Probabilistic Principal Component Analysis (PPCA) to reduce dimensions; 2) Removing all patients with mild OSA (AHI≥5 and <15) for training signal optimization; 3) Including medications as a features (using the first two digits of their GPI numbers); 4) Removing comorbidities or lab values from the training. Based on the exploratory data, we proceeded with the base model as our Full model, did not utilize PPCA, and did not remove the mild patients from the training dataset.

| **Any OSA** | | | | | | |
|---|---|---|---|---|---|---|
| **Model** | **Base** | **+Meds** | **+Meds; -Comorbid** | **-Labs** | **-Comorbid** | **-Mild** |
| HGB (PPCA) | 0.76 | 0.761 | 0.76 | 0.75 | 0.76 | |
| XGB (PPCA) | 0.76 | 0.7572 | 0.755 | 0.749 | 0.755 | 0.764 |
| RF (PPCA) | 0.753 | 0.756 | 0.754 | 0.747 | 0.754 | 0.757 |
| HGB (No PPCA) | 0.763 | 0.764 | 0.761 | 0.753 | 0.76 | 0.763 |
| XGB (No PPCA) | 0.761 | | | | 0.759 | 0.764 |
| RF (No PPCA) | 0.755 | | | | 0.754 | 0.756 |
| **Modsev OSA** | | | | | | |
| **Model** | **Base** | **+Meds** | **+Meds; -Comorbid** | **-Labs** | **-Comorbid** | **-Mild** |
| HGB (PPCA) | 0.74 | 0.742 | 0.74 | 0.734 | 0.74 | 0.744 |
| XGB (PPCA) | 0.743 | 0.745 | 0.743 | 0.735 | 0.741 | 0.746 |
| RF (PPCA) | 0.741 | 0.741 | 0.739 | 0.731 | 0.739 | 0.74 |
| HGB (No PPCA) | 0.732 | 0.745 | 0.741 | 0.736 | 0.742 | |
| XGB (No PPCA) | 0.745 | | | | 0.742 | 0.749 |
| RF (No PPCA) | 0.74 | | | | 0.738 | 0.74 |

**Supplement S3 – Model Search Spaces and Optimal Parameters**

**XGB · Full features · Any OSA (AHI ≥ 5)**

*BayesSearchCV · n_iter = 50*

| Parameter | Search Space | Optimal Parameters | Scale |
|---|---|---|---|
| n_estimators | 1,300–4,000 | 2062 | *integer* |
| max_depth | 6–10 | 9 | *integer* |
| learning_rate | 0.005–0.05 | 0.005147 | *log-uniform* |
| subsample | 0.60–0.95 | 0.6383 | *uniform* |
| colsample_bytree | 0.50–1.00 | 1 | *uniform* |
| colsample_bylevel | 0.40–0.80 | 0.4 | *uniform* |
| min_child_weight | 10–100 | 16 | *integer* |
| gamma | 0–2 | 1.347 | *uniform* |
| reg_alpha | 1e-4–1 | 0.3265 | *log-uniform* |
| reg_lambda | 1e-4–5 | 4.965 | *log-uniform* |

**XGB · Full features · Modsev OSA (AHI ≥ 15)**

*BayesSearchCV · n_iter = 50*

| Parameter | Search Space | Optimal Parameters | Scale |
|---|---|---|---|
| n_estimators | 1,300–4,000 | 4000 | *integer* |
| max_depth | 6–10 | 6 | *integer* |
| learning_rate | 0.003–0.02 | 0.005 | *log-uniform* |
| subsample | 0.60–0.95 | 0.6 | *uniform* |
| min_child_weight | 50–250 | 50 | *integer* |
| reg_alpha | 1e-4–1 | 1 | *log-uniform* |
| reg_lambda | 1e-4–15 | 15 | *log-uniform* |

**XGB · Minimal features · Any OSA (AHI ≥ 5)**

*BayesSearchCV · n_iter = 50*

| Parameter | Search Space | Optimal Parameters | Scale |
|---|---|---|---|
| n_estimators | 300–1,500 | 1229 | *integer* |
| max_depth | 3–8 | 3 | *integer* |
| learning_rate | 0.005–0.1 | 0.015242 | *log-uniform* |
| subsample | 0.60–0.95 | 0.6014 | *uniform* |
| min_child_weight | 30–300 | 32 | *integer* |
| colsample_bytree | 0.50–1.00 | 1 | *uniform* |

**XGB · Minimal features · Modsev OSA (AHI ≥ 15)**

*BayesSearchCV · n_iter = 50*

| Parameter | Search Space | Optimal Parameters | Scale |
|---|---|---|---|
| n_estimators | 300–1,500 | 1111 | *integer* |
| max_depth | 3–8 | 3 | *integer* |
| learning_rate | 0.005–0.1 | 0.016396 | *log-uniform* |
| subsample | 0.60–0.95 | 0.6 | *uniform* |
| min_child_weight | 30–300 | 30 | *integer* |
| colsample_bytree | 0.50–1.00 | 1 | *uniform* |

**RF · Full features · Any OSA (AHI ≥ 5)**

**RF · Full features · Modsev OSA (AHI ≥ 15)**

| | | | | | | | |
|---|---|---|---|---|---|---|---|
| *BayesSearchCV · n_iter = 50* | | | | *BayesSearchCV · n_iter = 50* | | | |
| **Parameter** | **Search Space** | **Optimal Parameters** | **Scale** | **Parameter** | **Search Space** | **Optimal Parameters** | **Scale** |
| n_estimators | 200–2,000 | 2000 | *integer* | n_estimators | 200–2,000 | 763 | *integer* |
| max_depth | 3–20 | 18 | *integer* | max_depth | 3–20 | 17 | *integer* |
| max_features | 0.10–1.00 | 0.5142 | *uniform* | max_features | 0.10–1.00 | 1 | *uniform* |
| min_samples_leaf | 1–50 | 1 | *integer* | min_samples_leaf | 1–50 | 1 | *integer* |
| min_samples_split | 2–50 | 2 | *integer* | min_samples_split | 2–50 | 15 | *integer* |
| max_samples | 0.50–1.00 | 0.5 | *uniform* | max_samples | 0.50–1.00 | 0.5 | *uniform* |
| ccp_alpha | 1e-4–1e-1 | 0.0001 | *log-uniform* | ccp_alpha | 1e-4–1e-1 | 0.0001 | *log-uniform* |
| **RF · Minimal features · Any OSA (AHI ≥ 5)** | | | | **RF · Minimal features · Modsev OSA (AHI ≥ 15)** | | | |
| *BayesSearchCV · n_iter = 30* | | | | *BayesSearchCV · n_iter = 30* | | | |
| **Parameter** | **Search Space** | **Optimal Parameters** | **Scale** | **Parameter** | **Search Space** | **Optimal Parameters** | **Scale** |
| n_estimators | 100–1,500 | 538 | *integer* | n_estimators | 100–1,500 | 581 | *integer* |
| max_depth | 3–20 | 17 | *integer* | max_depth | 3–20 | 10 | *integer* |
| min_samples_leaf | 10–200 | 120 | *integer* | min_samples_leaf | 10–200 | 35 | *integer* |
| min_samples_split | 2–100 | 29 | *integer* | min_samples_split | 2–100 | 95 | *integer* |
| max_features | 0.30–1.00 | 0.8189 | *uniform* | max_features | 0.30–1.00 | 0.4644 | *uniform* |
| max_samples | 0.50–1.00 | 0.8618 | *uniform* | max_samples | 0.50–1.00 | 0.9091 | *uniform* |
| ccp_alpha | 0.00–0.01 | 0.00003079 | *uniform* | ccp_alpha | 0.00–0.01 | 0.00004946 | *uniform* |
| **HGB · Full features · Any OSA (AHI ≥ 5)** | | | | **HGB · Full features · Modsev OSA (AHI ≥ 15)** | | | |
| *BayesSearchCV · n_iter = 50* | | | | *BayesSearchCV · n_iter = 50* | | | |
| **Parameter** | **Search Space** | **Optimal Parameters** | **Scale** | **Parameter** | **Search Space** | **Optimal Parameters** | **Scale** |
| max_iter | 1000–3,000 | 3000 | *integer* | max_iter | 1000–3,000 | 2719 | *integer* |
| max_depth | 6–10 | 10 | *integer* | max_depth | 6–10 | 10 | *integer* |
| learning_rate | 0.005–0.05 | 0.00631 | *log-uniform* | learning_rate | 0.005–0.05 | 0.006785 | *log-uniform* |
| max_features | 0.50–1.00 | 0.5 | *uniform* | max_features | 0.50–1.00 | 0.5 | *uniform* |
| min_samples_leaf | 10–100 | 100 | *integer* | min_samples_leaf | 10–100 | 100 | *integer* |

| | | | | | | | |
|---|---|---|---|---|---|---|---|
| l2_regularization | 1e-4–5 | 5 | *log-uniform* | l2_regularization | 1e-4–5 | 5 | *log-uniform* |
| max_bins | 64–255 | 255 | *integer* | max_bins | 64–255 | 255 | *integer* |
| **HGB · Minimal features · Any OSA (AHI ≥ 5)** | | | | **HGB · Minimal features · Modsev OSA (AHI ≥ 15)** | | | |
| *BayesSearchCV · n_iter = 50* | | | | *BayesSearchCV · n_iter = 50* | | | |
| **Parameter** | **Search Space** | **Optimal Parameters** | **Scale** | **Parameter** | **Search Space** | **Optimal Parameters** | **Scale** |
| max_iter | 300–2,000 | 1307 | *integer* | max_iter | 300–2,000 | 620 | *integer* |
| max_depth | 3–10 | 6 | *integer* | max_depth | 3–10 | 4 | *integer* |
| learning_rate | 0.005–0.1 | 0.034986 | *log-uniform* | learning_rate | 0.005–0.1 | 0.07839 | *log-uniform* |
| min_samples_leaf | 20–200 | 127 | *integer* | min_samples_leaf | 20–200 | 102 | *integer* |
| l2_regularization | 0.0–10.0 | 6.1708 | *uniform* | l2_regularization | 0.0–10.0 | 4.4483 | *uniform* |
| max_features | 0.30–1.00 | 0.6234 | *uniform* | max_features | 0.30–1.00 | 0.6033 | *uniform* |

#### Supplement S4 – SLIM Algorithm for OSA Screening

**Table 4**—SLIM scoring system (size 5, extractable features, FPR goal of 20%).

| PREDICT PATIENT HAS OSA IF TOTAL POINTS > 1 | | | |
|---|---|---|---|
| 1. Age ≥ 60 | 4 points | + | ________ |
| 2. Hypertension | 4 points | + | ________ |
| 3. BMI ≥ 30 | 2 points | + | ________ |
| 4. BMI ≥ 40 | 2 points | + | ________ |
| 5. Female | −6 points | − | ________ |
| **ADD POINTS FROM ROWS 1–5** | **TOTAL** | **=** | |

This model has a 10-CV mean TPR/FPR of 61.6%/20.8% and a training TPR/FPR of 62.0%/19.6%. BMI, body mass index; CV, cross-validation; OSA, obstructive sleep apnea; TPR, true positive rate; FPR, false positive rate; size refers to the number of features in the model.

SLIM is an algorithm for screening OSA utilizing 4 features that can be extractable from EHR. A SLIM score of >1 indicates risk for OSA.

Reference: Ustun B, Westover MB, Rudin C, Bianchi MT. Clinical Prediction Models for Sleep Apnea: The Importance of Medical History over Symptoms. J Clin Sleep Med. 2016 Feb;12(2):161-8. doi: 10.5664/jcsm.5476. PMID: 26350602; PMCID: PMC4751423.

**Supplement S5 – Cross-Validation Out-Of-Fold Results**

| **Full Model** | | | | | | |
|---|---|---|---|---|---|---|
| | **XGBoost (5+)** | **XGBoost (15+)** | **RF (5+)** | **RF (15+)** | **HGBC-MC (5+)** | **HGBC-MC (15+)** |
| **OOF Pooled AUC (95% CI)** | 0.791 (0.789-0.793) | 0.767 (0.765-0.769) | 0.781 (0.779-0.784) | 0.753 (0.751-0.755) | 0.790 (0.788-0.792) | 0.766 (0.764-0.768) |
| **Race** | | | | | | |
| Black (16368) | 0.781 (0.773-0.788) | 0.758 (0.751-0.765) | 0.771 (0.763-0.780) | 0.746 (0.738-0.753) | 0.780 (0.772-0.788) | 0.758 (0.751-0.765) |
| White (78771) | 0.784 (0.780-0.788) | 0.757 (0.754-0.761) | 0.776 (0.772-0.780) | 0.745 (0.742-0.749) | 0.783 (0.779-0.787) | 0.757 (0.754-0.760) |
| Asian (22332) | 0.793 (0.787-0.800) | 0.769 (0.762-0.775) | 0.779 (0.773-0.786) | 0.750 (0.743-0.756) | 0.793 (0.786-0.799) | 0.768 (0.761-0.774) |
| Hispanic (83446) | 0.797 (0.793-0.800) | 0.773 (0.769-0.776) | 0.790 (0.786-0.793) | 0.761 (0.758-0.764) | 0.796 (0.792-0.799) | 0.772 (0.769-0.775) |
| **Gender** | | | | | | |
| Male (120272) | 0.773 (0.770-0.776) | 0.746 (0.744-0.749) | 0.762 (0.759-0.765) | 0.730 (0.727-0.733) | 0.772 (0.768-0.775) | 0.746 (0.743-0.748) |
| Female (91907) | 0.783 (0.780-0.786) | 0.750 (0.747-0.753) | 0.772 (0.769-0.776) | 0.734 (0.731-0.738) | 0.782 (0.779-0.785) | 0.749 (0.746-0.752) |
| **Age** | | | | | | |
| 65 or Older (46198) | 0.673 (0.667-0.679) | 0.673 (0.668-0.678) | 0.665 (0.658-0.671) | 0.662 (0.657-0.667) | 0.671 (0.665-0.677) | 0.672 (0.667-0.677) |
| Under 65 (165990) | 0.810 (0.808-0.812) | 0.788 (0.786-0.790) | 0.800 (0.797-0.802) | 0.774 (0.771-0.776) | 0.809 (0.807-0.811) | 0.787 (0.785-0.790) |
| **Modality** | | | | | | |
| HSAT (210982) | 0.792 (0.789-0.794) | 0.767 (0.765-0.769) | 0.782 (0.780-0.784) | 0.753 (0.751-0.755) | 0.791 (0.788-0.793) | 0.766 (0.764-0.768) |
| PSG (1206) | 0.773 (0.747-0.799) | 0.783 (0.755-0.811) | 0.763 (0.736-0.789) | 0.758 (0.729-0.787) | 0.774 (0.749-0.800) | 0.783 (0.756-0.811) |

| Minimal Model | | | | | | |
|---|---|---|---|---|---|---|
| | XGBoost Min (5+) | XGBoost Min (15+) | RF Min (5+) | RF Min (15+) | HGBC Min (5+) | HGBC Min (15+) |
| **OOF Pooled AUC (95% CI)** | 0.766 (0.763-0.768) | 0.737 (0.735-0.739) | 0.763 (0.761-0.766) | 0.735 (0.733-0.737) | 0.765 (0.763-0.768) | 0.736 (0.734-0.738) |
| **Race** | | | | | | |
| Black (16368) | 0.754 (0.746-0.763) | 0.730 (0.722-0.738) | 0.752 (0.743-0.760) | 0.728 (0.720-0.735) | 0.754 (0.745-0.762) | 0.730 (0.722-0.737) |
| White (78771) | 0.759 (0.755-0.762) | 0.728 (0.724-0.731) | 0.757 (0.753-0.761) | 0.727 (0.723-0.730) | 0.758 (0.754-0.762) | 0.727 (0.724-0.731) |
| Asian (22332) | 0.760 (0.753-0.767) | 0.729 (0.723-0.736) | 0.752 (0.745-0.759) | 0.726 (0.720-0.733) | 0.759 (0.752-0.767) | 0.728 (0.722-0.735) |
| Hispanic (83446) | 0.774 (0.770-0.778) | 0.744 (0.741-0.747) | 0.773 (0.770-0.777) | 0.743 (0.740-0.747) | 0.774 (0.770-0.778) | 0.744 (0.740-0.747) |
| **Gender** | | | | | | |
| Male (120272) | 0.741 (0.738-0.745) | 0.709 (0.707-0.712) | 0.739 (0.735-0.742) | 0.708 (0.705-0.711) | 0.741 (0.737-0.744) | 0.709 (0.706-0.712) |
| Female (91907) | 0.758 (0.755-0.761) | 0.717 (0.714-0.720) | 0.755 (0.752-0.758) | 0.715 (0.712-0.718) | 0.758 (0.755-0.761) | 0.716 (0.713-0.720) |
| **Age** | | | | | | |
| 65 or Older (46198) | 0.653 (0.647-0.659) | 0.650 (0.645-0.655) | 0.652 (0.645-0.658) | 0.649 (0.644-0.654) | 0.652 (0.646-0.658) | 0.649 (0.644-0.654) |
| Under 65 (165990) | 0.783 (0.781-0.786) | 0.756 (0.754-0.759) | 0.781 (0.778-0.783) | 0.755 (0.753-0.757) | 0.783 (0.781-0.785) | 0.756 (0.754-0.759) |
| **Modality** | | | | | | |
| HSAT (210982) | 0.766 (0.764-0.768) | 0.737 (0.735-0.739) | 0.764 (0.761-0.766) | 0.735 (0.733-0.737) | 0.766 (0.763-0.768) | 0.736 (0.734-0.738) |
| PSG (1206) | 0.750 (0.723-0.776) | 0.752 (0.723-0.781) | 0.749 (0.722-0.776) | 0.751 (0.722-0.780) | 0.752 (0.725-0.779) | 0.753 (0.723-0.782) |

**Supplement S6 – Temporal and External Set Recalibrations.** Models underwent sigmoid recalibration on the temporal held-out set, even though they were already well calibrated. Recalibration may be necessary when applying to external datasets.

#### Temporal Set

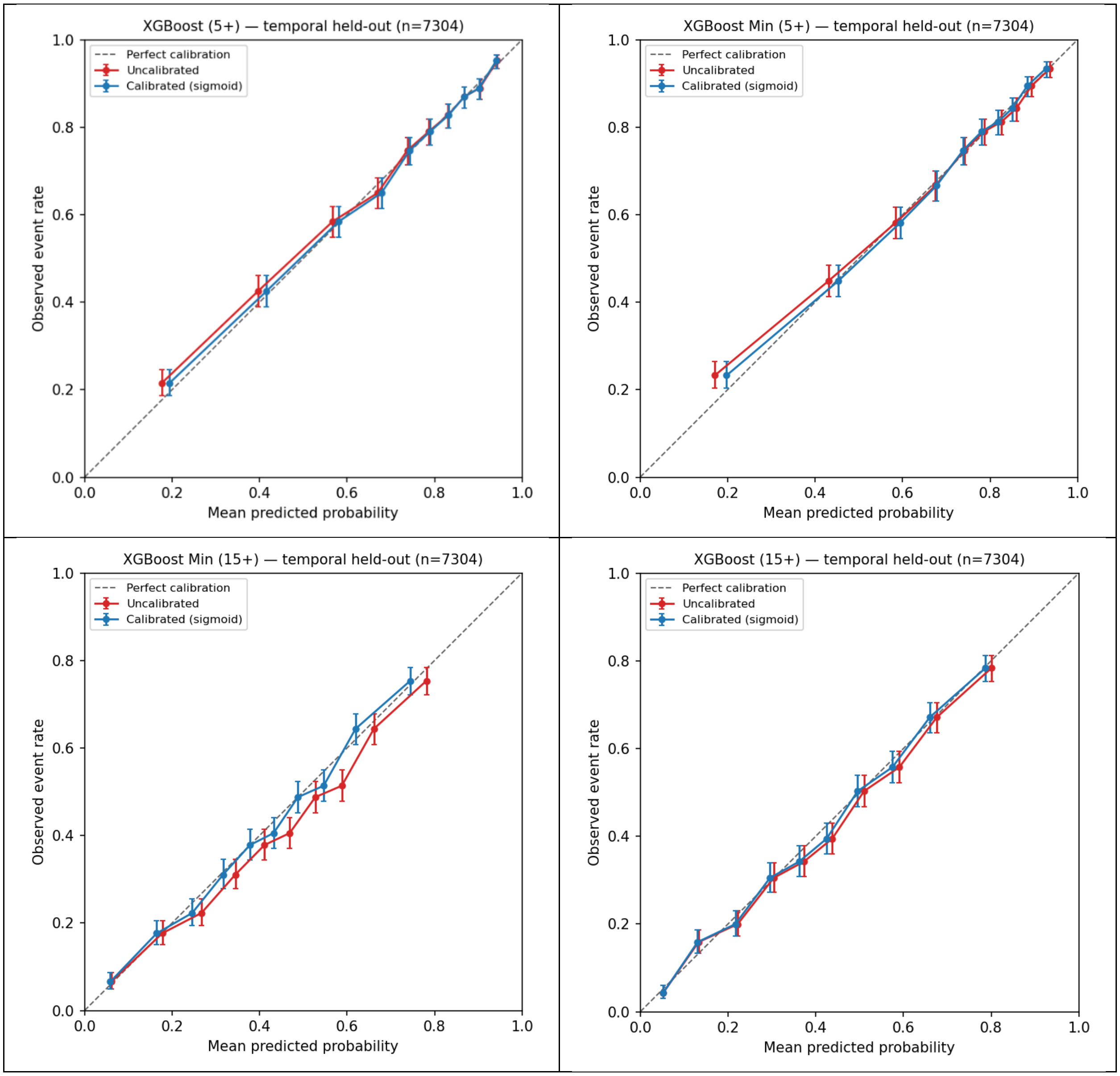

#### External Set

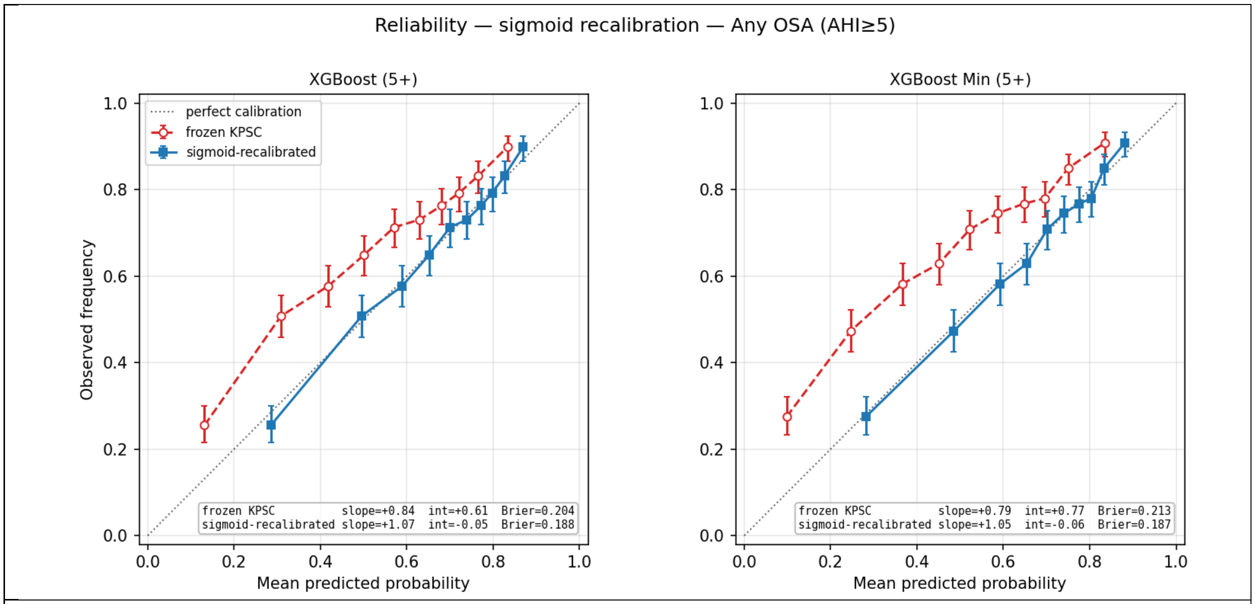

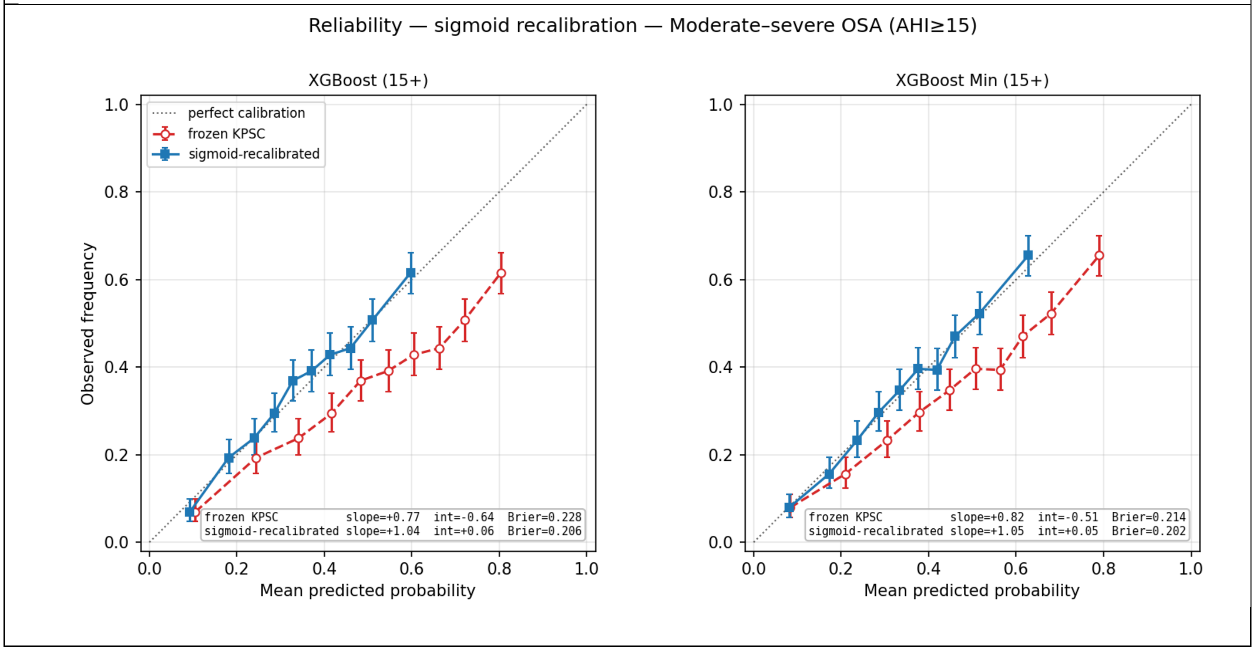

### Supplement S7 – XGBoost Feature Importance

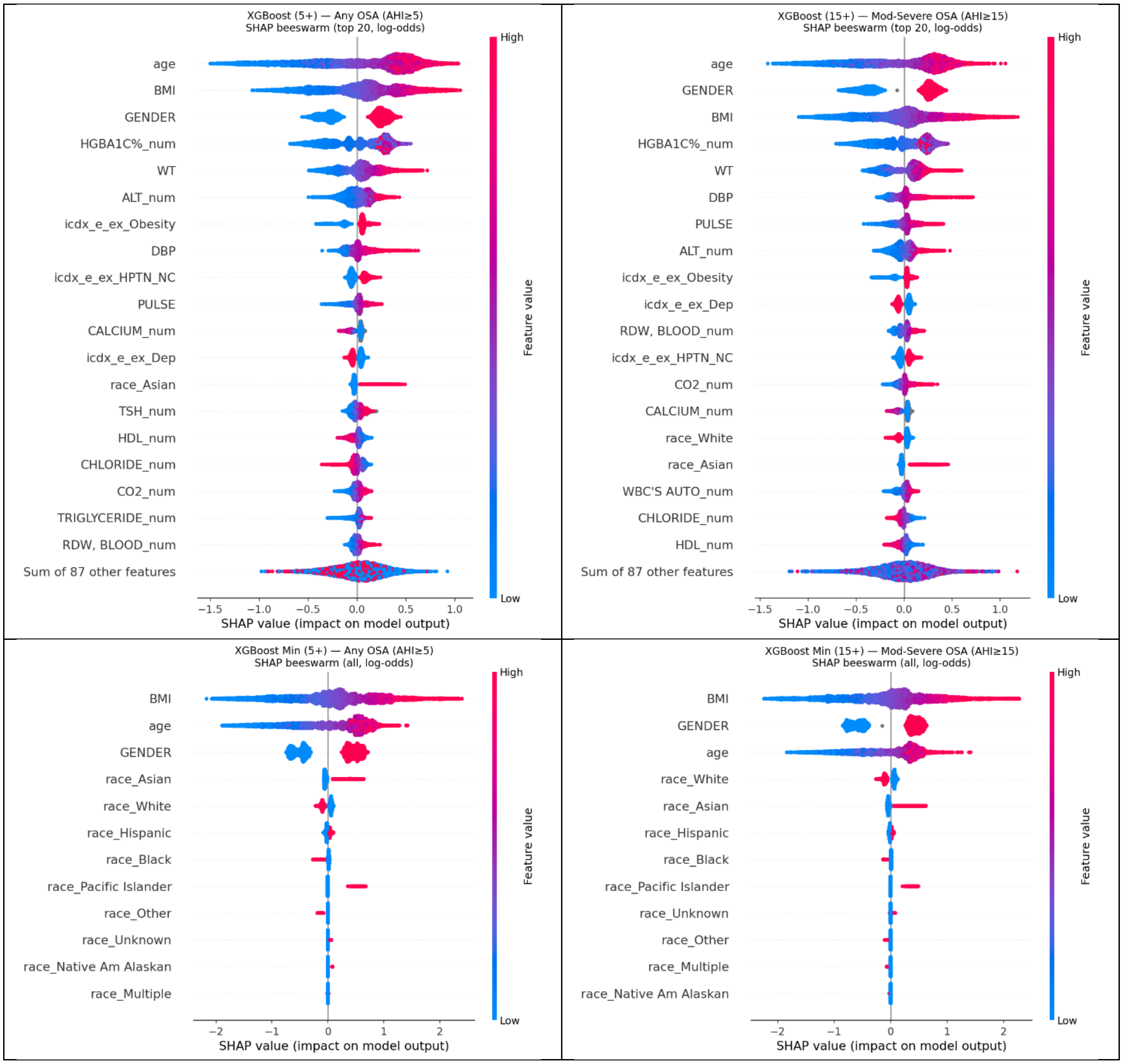

#### Supplement S8 – STOP-BANG/SLIM Comparison and Bayesian Risk-Updating Analysis Baseline Characteristics

| Feature | STOP-BANG/SLIM Comparison | Bayesian Analysis | STO LR Cohort | ESS LR Cohort |
|---|---|---|---|---|
| Total N | 1981 | 7286 | 29650 | 32486 |
| **DEMOGRAPHIC** | | | | |
| Age - mean (SD) | 54.6 (15.1) | 49.4 (15.3) | 49.7 (15.2) | 49.5 (15.1) |
| Gender - # (%) male | 1498 (75.6%) | 3884 (53.3%) | 15846 (53.4%) | 17406 (53.6%) |
| Race/Ethnicity - Asian - # (%) | 234 (11.8%) | 811 (11.1%) | 3293 (11.1%) | 3604 (11.1%) |
| Race/Ethnicity - Black - # (%) | 128 (6.5%) | 537 (7.4%) | 2184 (7.4%) | 2489 (7.7%) |
| Race/Ethnicity - Hispanic - # (%) | 558 (28.2%) | 2659 (36.5%) | 10829 (36.5%) | 11750 (36.2%) |
| Race/Ethnicity - Multiple - # (%) | 11 (0.6%) | 42 (0.6%) | 143 (0.5%) | 158 (0.5%) |
| Race/Ethnicity - Native Am Alaskan - # (%) | 4 (0.2%) | 15 (0.2%) | 51 (0.2%) | 57 (0.2%) |
| Race/Ethnicity - Other - # (%) | 44 (2.2%) | 148 (2.0%) | 504 (1.7%) | 575 (1.8%) |
| Race/Ethnicity - Pacific Islander - # (%) | 14 (0.7%) | 63 (0.9%) | 253 (0.9%) | 270 (0.8%) |
| Race/Ethnicity - Unknown - # (%) | 72 (3.6%) | 197 (2.7%) | 904 (3.0%) | 993 (3.1%) |
| Race/Ethnicity - White - # (%) | 916 (46.2%) | 2814 (38.6%) | 11489 (38.7%) | 12590 (38.8%) |
| **CLINICAL** | | | | |
| BMI - # (%) available | 1981 (100.0%) | 6015 (82.6%) | 24409 (82.3%) | 26739 (82.3%) |
| BMI - mean (SD) | 32.5 (7.4) | 33.9 (8.4) | 33.9 (8.5) | 33.9 (8.5) |
| AHI - mean (SD) | 21.0 (22.6) | 19.8 (23.2) | 20.4 (23.7) | 20.4 (23.8) |
| No OSA (AHI<5) - # (%) | 516 (26.0%) | 2365 (32.5%) | 9462 (31.9%) | 10517 (32.4%) |
| Mild OSA (AHI 5-<15) - # (%) | 579 (29.2%) | 1933 (26.5%) | 7805 (26.3%) | 8428 (25.9%) |
| Moderate-Severe OSA (AHI>=15) - # (%) | 886 (44.7%) | 2988 (41.0%) | 12383 (41.8%) | 13541 (41.7%) |
| **COMORBIDITIES** | | | | |
| Charlson Score (SD) | 1.9 (2.3) | 1.7 (2.2) | 1.8 (2.3) | 1.8 (2.3) |
| Hypertension - # (%) | 995 (50.2%) | 3380 (46.4%) | 13926 (47.0%) | 15113 (46.5%) |
| Depression - # (%) | 830 (41.9%) | 3633 (49.9%) | 14526 (49.0%) | 15964 (49.1%) |
| Diabetes - # (%) | 453 (22.9%) | 1545 (21.2%) | 6642 (22.4%) | 7243 (22.3%) |
| Heart Failure - # (%) | 138 (7.0%) | 432 (5.9%) | 1929 (6.5%) | 2095 (6.4%) |
| Cerebrovascular Disease - # (%) | 149 (7.5%) | 423 (5.8%) | 1910 (6.4%) | 2064 (6.4%) |
| Renal Disease - # (%) | 226 (11.4%) | 748 (10.3%) | 3165 (10.7%) | 3436 (10.6%) |
| **LABORATORY** | | | | |
| HgbA1c - mean (SD) | 5.9 (0.9) | 5.9 (0.9) | 5.9 (0.9) | 5.9 (0.9) |
| HgbA1c - # (%) available | 1923 (97.1%) | 7011 (96.2%) | 28727 (96.9%) | 31439 (96.8%) |
| Glucose (fasting) - mean (SD) | 102.3 (29.4) | 99.8 (26.9) | 100.8 (30.7) | 100.9 (31.1) |
| Glucose (fasting) - # (%) available | 610 (30.8%) | 2091 (28.7%) | 8656 (29.2%) | 9560 (29.4%) |
| Cholesterol (total) - mean (SD) | 177.6 (42.7) | 181.5 (42.3) | 181.8 (41.2) | 182.0 (41.2) |
| Cholesterol (total) - # (%) available | 1923 (97.1%) | 6983 (95.8%) | 28618 (96.5%) | 31312 (96.4%) |

**Supplement S9 – Comparing Performance of XGBoost Models on AHI4% and AHI3%.** This exploratory analysis revealed similar AUC when applying the KPSC models to a KUMC cohort of patients with both AHI4% and AHI3% derived from the same polysomnography study.

| AHI3% vs AHI4% | | | | | | |
|---|---|---|---|---|---|---|
| **model** | **target** | **n** | **Prevalence 3%** | **Prevalence 4%** | **AUC 3%** | **AUC 4%** |
| XGBoost Full | Any OSA | 7998 | 0.816 | 0.654 | 0.747 | 0.735 |
| XGBoost Min | Any OSA | 7998 | 0.816 | 0.654 | 0.758 | 0.733 |
| XGBoost Full | Mod-Severe OSA | 7998 | 0.513 | 0.336 | 0.692 | 0.710 |
| XGBoost Min | Mod-Severe OSA | 7998 | 0.513 | 0.336 | 0.710 | 0.715 |

**Supplement S10 – Classification Performance Metrics.** KPSC metrics were calculated based on Youden's J-determined optimal threshold and when sensitivity was fixed at 80%. KUMC metrics were first calculated using KPSC thresholds; optimal thresholds were then recalculated using Youden's J on KUMC's training cohort. Sens = Sensitivity, Spec = Specificity, PPV = Positive Predictive Value, NPV = Negative Predictive Value.

| | | KPSC Sensitivity Fixed to 0.8 | | | | | KPSC Optimal Threshold | | | | | KUMC Optimal Threshold | | | | |
|---|---|---|---|---|---|---|---|---|---|---|---|---|---|---|---|---|
| **Model** | **Site** | **Threshold** | **Sens** | **Spec** | **PPV** | **NPV** | **Threshold** | **Sens** | **Spec** | **PPV** | **NPV** | **Threshold** | **Sens** | **Spec** | **PPV** | **NPV** |
| XGB Full (Any-OSA) | KPSC | 0.696 | 80% | 63% | 85% | 54% | 0.742 | 74% | 69% | 87% | 50% | | | | | |
| XGB Min (Any-OSA) | | 0.693 | 80% | 58% | 84% | 52% | 0.719 | 76% | 63% | 85% | 50% | | | | | |
| XGB Full (Modsev-OSA) | | 0.407 | 80% | 58% | 62% | 77% | 0.457 | 74% | 66% | 65% | 74% | | | | | |
| XGB Min (Modsev-OSA) | | 0.408 | 80% | 53% | 59% | 75% | 0.471 | 69% | 65% | 63% | 71% | | | | | |
| XGB Full (Any-OSA) | KUMC | 0.696 | 66% | 64% | 79% | 48% | 0.742 | 54% | 75% | 81% | 44% | 0.696 | 66% | 64% | 79% | 48% |
| XGB Min (Any-OSA) | | 0.693 | 68% | 65% | 80% | 49% | 0.719 | 61% | 70% | 81% | 47% | 0.701 | 66% | 66% | 80% | 49% |
| XGB Full (Modsev-OSA) | | 0.407 | 52% | 73% | 51% | 73% | 0.457 | 39% | 82% | 54% | 71% | 0.377 | 60% | 66% | 49% | 75% |
| XGB Min (Modsev-OSA) | | 0.408 | 55% | 72% | 52% | 74% | 0.471 | 37% | 86% | 59% | 71% | 0.352 | 70% | 59% | 48% | 78% |

**Supplement S11 – Sample Warm-Start Fine-Tuning Code for XGBoost Transfer Learning.** Our code was similar to this sample code found at: https://xgboost.readthedocs.io/en/stable/python/examples/continuation.html

```
import os
import pickle
import tempfile

from sklearn.datasets import HYPERLINK "https://scikit-learn.org/stable/modules/generated/sklearn.datasets.load_breast_cancer.html" \l "sklearn.datasets.load_breast_cancer" \o "sklearn.datasets.load_breast_cancer"

import xgboost

def training_continuation(tmpdir: str, use_pickle: bool) -> None:
    """Basic training continuation."""
    # Train 128 iterations in 1 session
    X, y = HYPERLINK "https://scikit-learn.org/stable/modules/generated/sklearn.datasets.load_breast_cancer.html" \l "sklearn.datasets.load_breast_cancer" \o "sklearn.datasets.load_breast_cancer"(return_X_y=True)
    clf = xgboost.XGBClassifier(n_estimators=128, eval_metric="logloss")
    clf.fit(X, y, eval_set=[(X, y)])
    print("Total boosted rounds:", clf.get_booster().num_boosted_rounds())

    # Train 128 iterations in 2 sessions, with the first one runs for 32 iterations and
    # the second one runs for 96 iterations
    clf = xgboost.XGBClassifier(n_estimators=32, eval_metric="logloss")
    clf.fit(X, y, eval_set=[(X, y)])
    assert clf.get_booster().num_boosted_rounds() == 32

    # load back the model, this could be a checkpoint
    if use_pickle:
        path = HYPERLINK "https://docs.python.org/3.12/library/os.path.html" \l "os.path.join" \o "os.path.join"(tmpdir, "model-first-32.pkl")
        with open(path, "wb") as fd:
            HYPERLINK "https://docs.python.org/3.12/library/pickle.html" \l "pickle.dump" \o "pickle.dump"(clf, fd)
        with open(path, "rb") as fd:
            loaded = HYPERLINK "https://docs.python.org/3.12/library/pickle.html" \l "pickle.load" \o "pickle.load"(fd)
    else:
        path = HYPERLINK "https://docs.python.org/3.12/library/os.path.html" \l "os.path.join" \o "os.path.join"(tmpdir, "model-first-32.json")
        clf.save_model(path)
        loaded = xgboost.XGBClassifier()
        loaded.load_model(path)

    clf = xgboost.XGBClassifier(n_estimators=128 - 32, eval_metric="logloss")
    clf.fit(X, y, eval_set=[(X, y)], xgb_model=loaded)

    print("Total boosted rounds:", clf.get_booster().num_boosted_rounds())

    assert clf.get_booster().num_boosted_rounds() == 128

def training_continuation_early_stop(tmpdir: str, use_pickle: bool) -> None:
    """Training continuation with early stopping."""
    early_stopping_rounds = 5
    early_stop = xgboost.callback.EarlyStopping(
        rounds=early_stopping_rounds, save_best=True
    )
    n_estimators = 512

    X, y = HYPERLINK "https://scikit-learn.org/stable/modules/generated/sklearn.datasets.load_breast_cancer.html" \l "sklearn.datasets.load_breast_cancer" \o "sklearn.datasets.load_breast_cancer"(return_X_y=True)
    clf = xgboost.XGBClassifier(
        n_estimators=n_estimators, eval_metric="logloss", callbacks=[early_stop]
    )
```

```
    clf.fit(X, y, eval_set=[(X, y)])
    print("Total boosted rounds:", clf.get_booster().num_boosted_rounds())
    best = clf.best_iteration

    # Train 512 iterations in 2 sessions, with the first one runs for 128 iterations and
    # the second one runs until early stop.
    clf = xgboost.XGBClassifier(
        n_estimators=128, eval_metric="logloss", callbacks=[early_stop]
    )
    # Reinitialize the early stop callback
    early_stop = xgboost.callback.EarlyStopping(
        rounds=early_stopping_rounds, save_best=True
    )
    clf.set_params(callbacks=[early_stop])
    clf.fit(X, y, eval_set=[(X, y)])
    assert clf.get_booster().num_boosted_rounds() == 128

    # load back the model, this could be a checkpoint
    if use_pickle:
        path = HYPERLINK "https://docs.python.org/3.12/library/os.path.html" \l "os.path.join" \o "os.path.join"(tmpdir, "model-first-128.pkl")
        with open(path, "wb") as fd:
            HYPERLINK "https://docs.python.org/3.12/library/pickle.html" \l "pickle.dump" \o "pickle.dump"(clf, fd)
        with open(path, "rb") as fd:
            loaded = HYPERLINK "https://docs.python.org/3.12/library/pickle.html" \l "pickle.load" \o "pickle.load"(fd)
    else:
        path = HYPERLINK "https://docs.python.org/3.12/library/os.path.html" \l "os.path.join" \o "os.path.join"(tmpdir, "model-first-128.json")
        clf.save_model(path)
        loaded = xgboost.XGBClassifier()
        loaded.load_model(path)

    early_stop = xgboost.callback.EarlyStopping(
        rounds=early_stopping_rounds, save_best=True
    )
    clf = xgboost.XGBClassifier(
        n_estimators=n_estimators - 128, eval_metric="logloss", callbacks=[early_stop]
    )
    clf.fit(
        X,
        y,
        eval_set=[(X, y)],
        xgb_model=loaded,
    )

    print("Total boosted rounds:", clf.get_booster().num_boosted_rounds())
    assert clf.best_iteration == best

if __name__ == "__main__":
    with HYPERLINK "https://docs.python.org/3.12/library/tempfile.html" \l "tempfile.TemporaryDirectory" \o "tempfile.TemporaryDirectory"() as tmpdir:
        training_continuation_early_stop(tmpdir, False)
        training_continuation_early_stop(tmpdir, True)

        training_continuation(tmpdir, True)
        training_continuation(tmpdir, False)
```
